# CD1c Dendritic Cells are depleted and accompanied by new HLA-DR^hi^ Phenotypes in Rheumatoid Arthritis Blood

**DOI:** 10.1101/2024.03.13.24304213

**Authors:** Christian Geier, Haani Qudsi, Jihad Ben Gabr, Robert Winchester, Andras Perl

**Author notes:** Corresponding author: Andras Perl, MD PhD Division of Rheumatology & Clinical Immunology SUNY Upstate Medical University. Funding and Disclosures: Grant funding for the work reported in this manuscript was provided by the Ines Mandl Research Foundation and SUNY Upstate Medical University (Department of Medicine); commercial sources were not involved. The authors have no conflicts of interest to report.

## Abstract

Rheumatoid arthritis (RA) in an autoimmune disease that leads to inflammation of synovial joints and other organs. Many RA patients ‘share’ a common peptide sequence within the HLA-DR (DR) molecule expressed on antigen-presenting cells (APC), suggesting that DR^hi^ cells are important in RA. Here, we use DR^hi^ to broadly define and immunophenotype RA APC, including potential APC not meeting standard definitions for lymphocytes, monocytes, dendritic cells (DC) from RA patients and healthy controls (HC). We measured mean fluorescence intensities (MFI) of molecules associated with DC (CD141, CD1c, CD11c, CD123, CD303), monocytes (CD14, CD16); granulocytic markers (CD15, CCR3), co-stimulatory molecules and chemokine receptors. DC2 (CD1c^+^) showed higher CD56, CD86, CD275, and CCR7 in RA. DC2 frequencies were much lower in RA: 3.2% of DR^hi^ [IQR 2.41 to 4.46] in RA vs. 6.9% [IQR 3.96 to 9.08] in HC; p=0.005. CD15 was increased in all RA APC subsets (p<0.01). A distinct CD15^+^CD16^+^ population appeared in RA, representing 1.5% of leukocytes [IQR 0.68 to 3.32] (vs 0.1% in HC [IQR 0.08 to 0.46]; p<0.001) and contributed a mean of 2.34% to overall DR^hi^. The CD15^+^CD16^+^ subset was CD303^+^, CD83^+^ and CD275^+^ with much less CD123 relative to reference plasmacytoid DC (p<0.01). In conclusion, APC alterations in RA include depletion of DC2 and increased CD15. Moreover, the APC (DR^hi^) compartment in RA contains cells with shared dendritic cell and granulocytic features; this phenotype suggests these apparent APC may participate in the pathophysiology of rheumatoid arthritis via the presentation of self-antigen(s) to CD4^+^ T lymphocytes.

## INTRODUCTION

In Rheumatoid arthritis (RA) aberrant lymphocytes can damage synovial joints and other organs (1). Antigen-presenting cells (APC) can activate lymphocytes and are considered critical to initiate immune responses (2). The profound association of the HLA-DR (DR) ‘Shared Epitope’ (SE) motif with rheumatoid arthritis (3,4) led us to reason that DR^hi^ APC are important in RA and that RA peripheral blood mononuclear cells (PBMC) may contain APC of pathophysiologic relevance beyond those meeting standard definitions of B lymphocytes and monocytes. We hypothesized that compared with healthy controls (HC) the blood of RA patients contains APC with increased inflammatory potential (↑ HLA-DR expression, co-stimulatory phenotype; impaired inhibitory marker expression) and chemokine receptor expression.

## MATERIALS AND METHODS

### Collection and isolation of peripheral blood mononuclear cells

We collected heparinized whole blood from RA patients meeting the 2010 ACR/EULAR classification criteria (5) and healthy control volunteers by venipuncture. Collection was approved by the institutional review board of SUNY Upstate Medical University (Syracuse, USA). Peripheral blood mononuclear cells (PBMCs) were isolated by Ficoll density gradient separation on the day of collection. PBMCs from the resulting monolayer were carefully aspirated by pipetting and washed twice in phosphate-buffered saline (PBS) with centrifugation at 350G for 7 minutes at room temperature, then resuspended in heat-inactivated fetal bovine serum (FBS) or, for some experiments, in RPMI-1640 medium supplemented with 10% FBS. We cryopreserved PBMCs by dropwise addition of a mixture of 10% dimethyl sulfoxide (DMSO) and 90% FBS to gradually achieve a final concentration of 5% DMSO. Cells were then cooled in a MrFrosty^TM^ freezing container (ThermoFisher, Waltham, USA) at a rate of approximately -1°C per minute to -80°C and — if timely analysis was feasible — kept at -80°C or, when necessary, transferred into the vapor phase of liquid nitrogen (LN2) for long-term storage.

### Recruitment of RA patients and healthy control donors

We collected basic demographic and clinical information including age, gender, smoking status, rheumatoid factor (RF) and anti-cyclic citrullinated peptide (CCP) antibody status (Supplementary Table 1). We recruited healthy volunteers for collection of peripheral blood and basic demographic information. We screened RA donors for patients for severe RA (polyarticular synovitis on exam) and designated three patients ‘index patients’ that were analyzed in detail to create working definitions for potentially antigen-presenting cells.

### Technical reference samples from healthy blood donors

In addition to biological controls, for some experiments, we used reference single healthy donor samples. We purchased buffy coats from healthy blood donors (New York Blood Center, New York, USA). PBMCs from buffy coats were isolated by Ficoll density gradient separation as described above but were centrifuged and washed three times in PBS at room temperature. Aliquots of single donor PBMCs were stored in LN2 for use as technical reference controls over the course of the study.

### Spectral flow cytometry Panel design

The spectral cytometry panel for antigen-presenting cells (Supplementary Table 2) consisted of — from BD Biosciences (Franklin Lakes, USA): CD16-BUV496 (clone 3G8), CD56-BUV737 (NCAM16.2), CD45RA-BUV395 (HI100), HLA-DR-V500 (G46-6), CD141-BB515 (1A4). BioLegend (San Diego, USA): CD123-BV510 (6H6), CCR7- BV421 (G043H7), CD19-PerCP-Cy5.5 (HIB19), CD14-SparkBlue550 (63D3), CD45- PerCP (2D1), CCR2-PE-Cy7 (K036C2), CD303-APC-Fire750 (201A), CD1c-AF647 (L161), CD83-PE-Cy5 (HB15e), CD86-BV711 (IT2.2), CD155-PE/Dazzle594 (SKII.4); from ThermoFisher: CD3-PerCP-Cy5.5 (SK7), CD11c-eFluor450 (3.9) and viability staining: Live/Dead Fix Blue or Propidium iodide (PI). For some experiments, the following additional reagents were used: CD15-BV605 (W6D3), CCR3-BUV805 (5E8), CD19-SparkNIR 685 (HIB19).

Selection of fluorochromes was informed by a previously validated spectral cytometry panel design (6) that we modified for our purposes. During panel development we calculated staining indices to inform optimal concentrations and staining conditions. For all reagents, we confirmed robust discrimination of fluorochrome/antibody by signal to noise ratio in single stain and multicolor “cocktail” staining pilot experiments. For most reagents, 1 μL of conjugate per sample was selected as the most suitable and was added; for the following reagents 2 μL were added: CCR7-BV421 (G043H7), CD11c- eFluor450 (3.9), CD19-PerCP-Cy5.5 (HIB19), CD155-PE/Dazzle594 (SKII.4).

### Sample preparation for flow cytometry

Spectral flow cytometry of RA and healthy donor samples was performed in an interleaved fashion, in batches of four to six samples, and processed in parallel. PBMC from two samples at a time were retrieved from -80°C or LN2 storage and rapidly thawed in a 37°C water bath, followed by dropwise addition of warm complete RPMI. This was repeated either once (when four samples were analyzed), or twice for six samples. PBMCs were then washed in complete RPMI and centrifuged at 350G for 7 minutes at room temperature. Cell viability following thawing was assessed by trypan blue exclusion and typically exceeded 90%.

A cocktail of fluorochrome/antibody conjugates was prepared immediately prior to staining. Following the addition of the antibody cocktail, PBMC were incubated for 30 minutes at room temperature in the dark in a total staining volume of approximately 50 µl. PBS supplemented with 0.5% or 1% bovine serum albumin (BSA) was used as staining buffer (FACS buffer). Based on the results of optimization experiments, the stained cells were washed once in FACS buffer and resuspended in 125 µL of FACS buffer. For viability staining, we used Live/Dead Blue (ThermoFisher) according to the instructions provided by the manufacturer, or alternatively, added PI immediately prior to acquisition.

### Data acquisition

Flow cytometry experiments were conducted on a Cytek Aurora® 5-laser spectral cytometer (Cytek Biosciences, Fremont, USA) with a laser configuration as recommended by the manufacturer.

### Bi-axial gating strategy

FCS Express Versions 6 and 7 (DeNovo Software, Pasadena, USA) and SpectroFlo Version 3.0.3 (Cytek Biosciences) were used for bi-axial gating. To identify potential new antigen-presenting cells, expressing marker combinations not conforming to existing APC definitions, we gated all HLA-DR positive cells lacking lymphoid markers. (Supplementary Figure 1). Based on findings of increased CD15 in the reference population we used a revised gating strategy to quantify and characterize granulocytic cells, including those with APC potential (Supplementary Figure 2).

### T-distributed stochastic neighbor embedding

For dimensionality reduction we applied a t-distributed stochastic neighbor embedding (t-SNE) algorithm. We used the Barnes-Hut implementation in FCS Express Version 7 (DeNovo Software, Pasadena, USA) with the following parameters: perplexity 100, iterations 500, seed 6; resolution for the resulting plots was 512x512 with smoothing 1. We annotated the resulting embeddings in FCS Express for those clusters corresponding to ‘established’ non-lymphoid APC (monocyte and DC subsets) and those that did not correspond to these gating paradigms.

### Statistical analysis

We used two-sided t-tests or Kruskal-Wallis testing to determine significant differences between RA and healthy individuals for APC subsets; a threshold of p<0.05 was considered significant. We preplanned comparisons of APC with plausible biological significance (HLA-DR^+^) between RA and healthy donors and reported all comparisons.

## RESULTS

Since dendritic cells (DC) are considered the most potent representatives of the system of antigen-presenting cells (APC) that initiate immune responses (7), we focused our initial attention on this APC lineage. We used spectral cytometry to characterize DC (CD141^+^ DC1, CD1c^+^ DC2, CD123^+^CD303^+^ pDC) in RA and healthy control donors (Supplementary Table 1)

### Dendritic Cells exhibit an inflammatory phenotype in RA blood

DC2 (CD1c^+^) — the most abundant DC subset (8) and considered to be the most potent APC (9) — displayed the most profound phenotypic alterations of the DC subsets. Compared with DC2 from healthy controls, RA DC2 differed by higher mean fluorescence intensity (MFI) of CD56 (Figure 1A and Supplementary Table 3). Furthermore, consistent with an inflammatory state, RA DC2 displayed higher MFIs of the co-stimulatory molecules CD86 and CD275 (ICOS-L) as well as C-C chemokine receptor type 7 (CCR7) (Figure 1B-D and Supplementary Table 3). Similarly, DC1 (CD141^+^) showed significant increases in CCR7 and CD275 (ICOS-L) in RA (Supplementary Table 4 and Supplementary Figure 3D, V); CD86 did not differ (Supplementary Table 4 and Supplementary Figure 3Q). In plasmacytoid dendritic cells (CD303^+^CD123^+^ pDC), changes in RA were largely restricted to increased CD56 and low-level expression of CD83 (Supplementary Table 5 and Supplementary Figure 5O). Despite their inflammatory (co-stimulatory) phenotype, RA monocytes and dendritic cells lacked the anticipated increased HLA-DR expression (MFI); whereas in reference RA B cells (CD19^+^), HLA-DR was increased (Supplementary Figure 6X), as expected.

**Figure 1.**
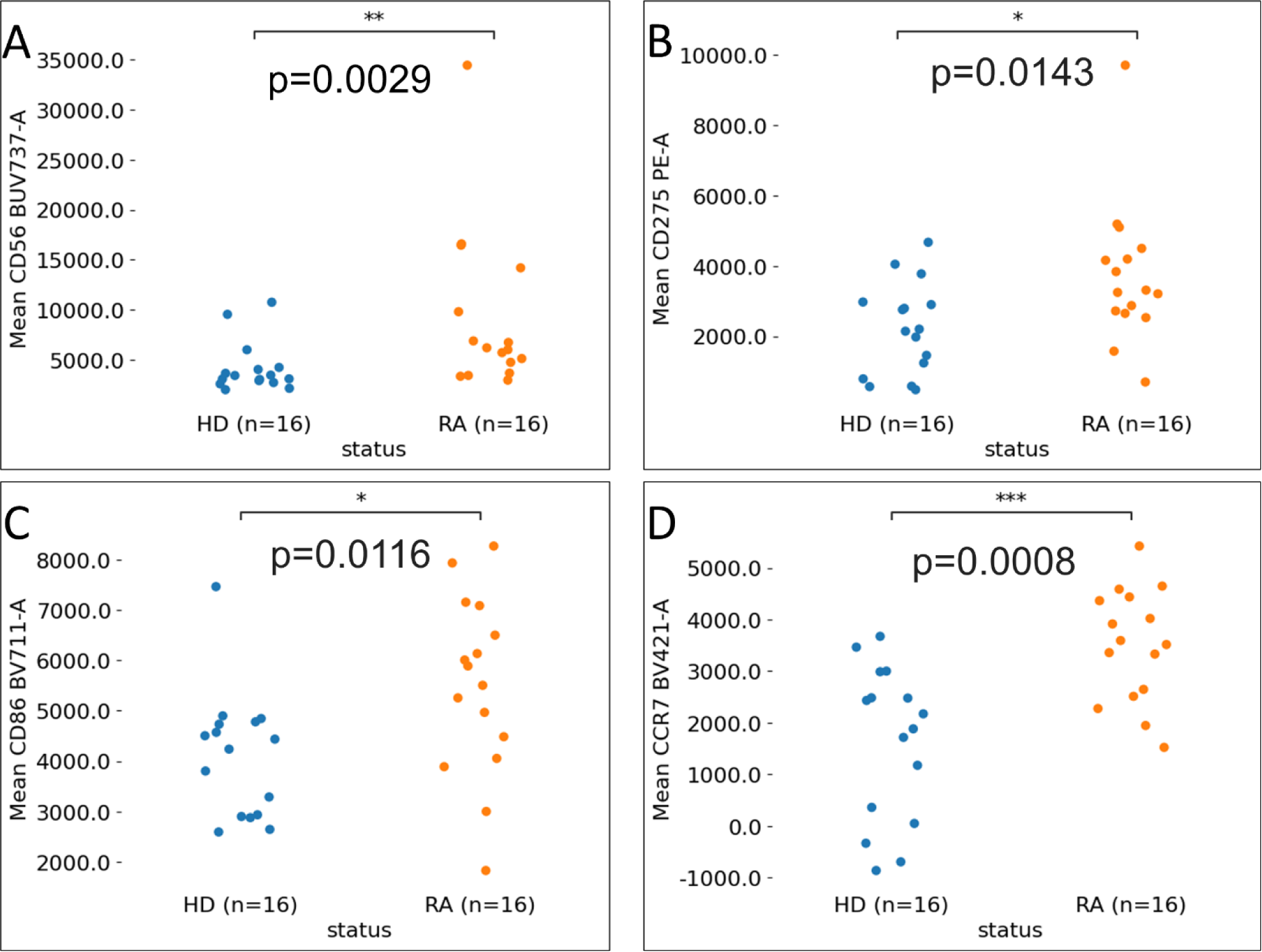
CD1c Dendritic Cell (DC2) phenotypes in healthy control donors (HD, blue) and rheumatoid arthritis patients (RA, orange). RA DC2 are characterized by higher CD56 (**A**), the co-stimulatory molecules CD86 and CD275 (ICOS-L) (**B, C**) and C-C chemokine receptor type 7 (**D**). Mean fluorescence intensities (MFI) measured by spectral cytometry.

### The DR^hi^ compartment of RA blood lacks CD1c^+^ dendritic cells (DC2)

We then quantified the contribution of dendritic cells to DR^hi^ in healthy individuals and RA donors. Surprisingly, in RA blood, CD1c^+^ dendritic cells (DC2) were approximately halved (3.21% vs. 6.87%; p=0.005; Figure 2A). Contributions of CD141^+^ dendritic cells (DC1) and plasmacytoid DC to the DR^hi^ APC compartment were numerically lower in RA but did not differ statistically from healthy controls (Figure 2B-C). Consistent with earlier findings (10), intermediate monocytes (CD14^+^CD16^+^) were higher in RA (14.25% [IQR 7.70 to 17.98] vs. 4.74% [1.21 to 6.64]; p=0.0037; Supplementary Table 8). Proportions of classical (CD14^+^CD16^-^) and non-classical (CD14^lo^CD16^+^) monocytes did not differ (Supplementary Table 8).

**Figure 2.**
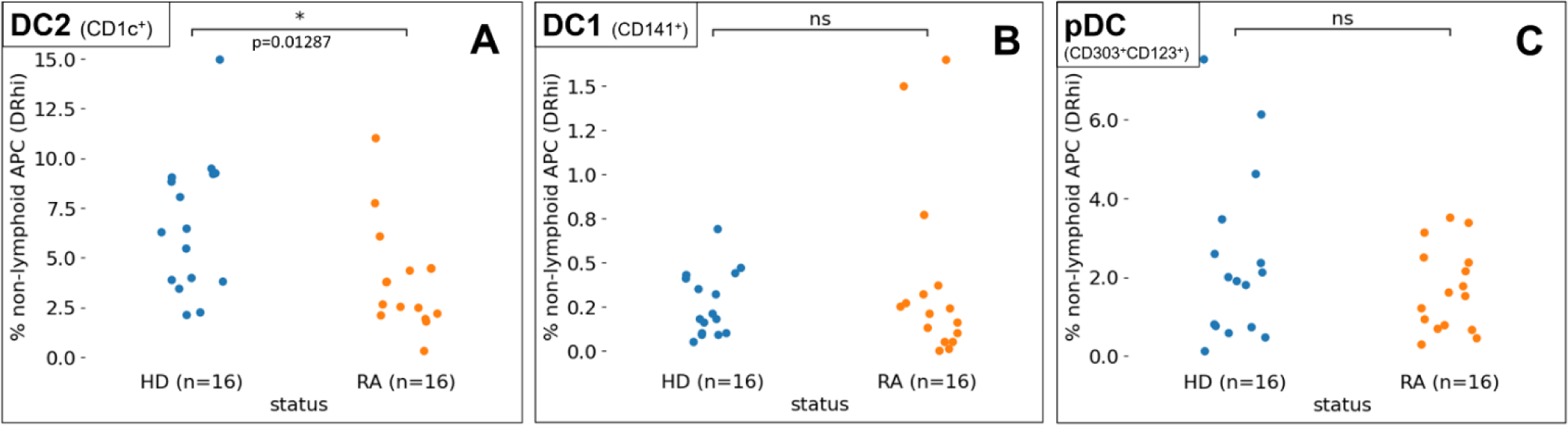
Quantification of Dendritic cell (DC) subsets by spectral cytometry from healthy control donors (HD, blue) and patients with rheumatoid arthritis (RA, orange), in % of non-lymphoid APC (DR^hi^). DC2 (CD1c^+^ dendritic cells) are decreased in RA patients (p=0.01287) (**A**) The proportions of DC1 (CD141^+^ dendritic cells) and plasmacytoid DC (CD303^+^CD123^+^) did not differ (p>0.05) (**B, C**)

### The granulocyte-associated molecule CD15 is markedly increased in RA APC

Given the unexpected lack of an HLA-DR increase in monocytes and DC and the observed apparent shift in the DR^hi^ compartment away from CD1c^+^ dendritic cells (DC2) in RA we decided to re-analyze the DR^hi^ compartment for additional lineage shifts. We quantified MFIs for CD15 and CCR3, molecules generally associated with mono/granulocytic myeloid lineages (11,12). CD15, virtually absent in healthy donors, was dramatically increased in all dendritic cell and monocyte subsets in RA (Figure 3A- F; p<0.01). In contrast, CCR3 intensities did not differ in RA (Supplementary Figures 2C-6C; 8C), except for a modest increase in intermediate monocytes (Supplementary Figure 7C). This apparent CD15 signature within DR^hi^ RA DC prompted us to carefully re-analyze the DR^hi^ compartment for additional CD15-positive phenotypes.

**Figure 3.**
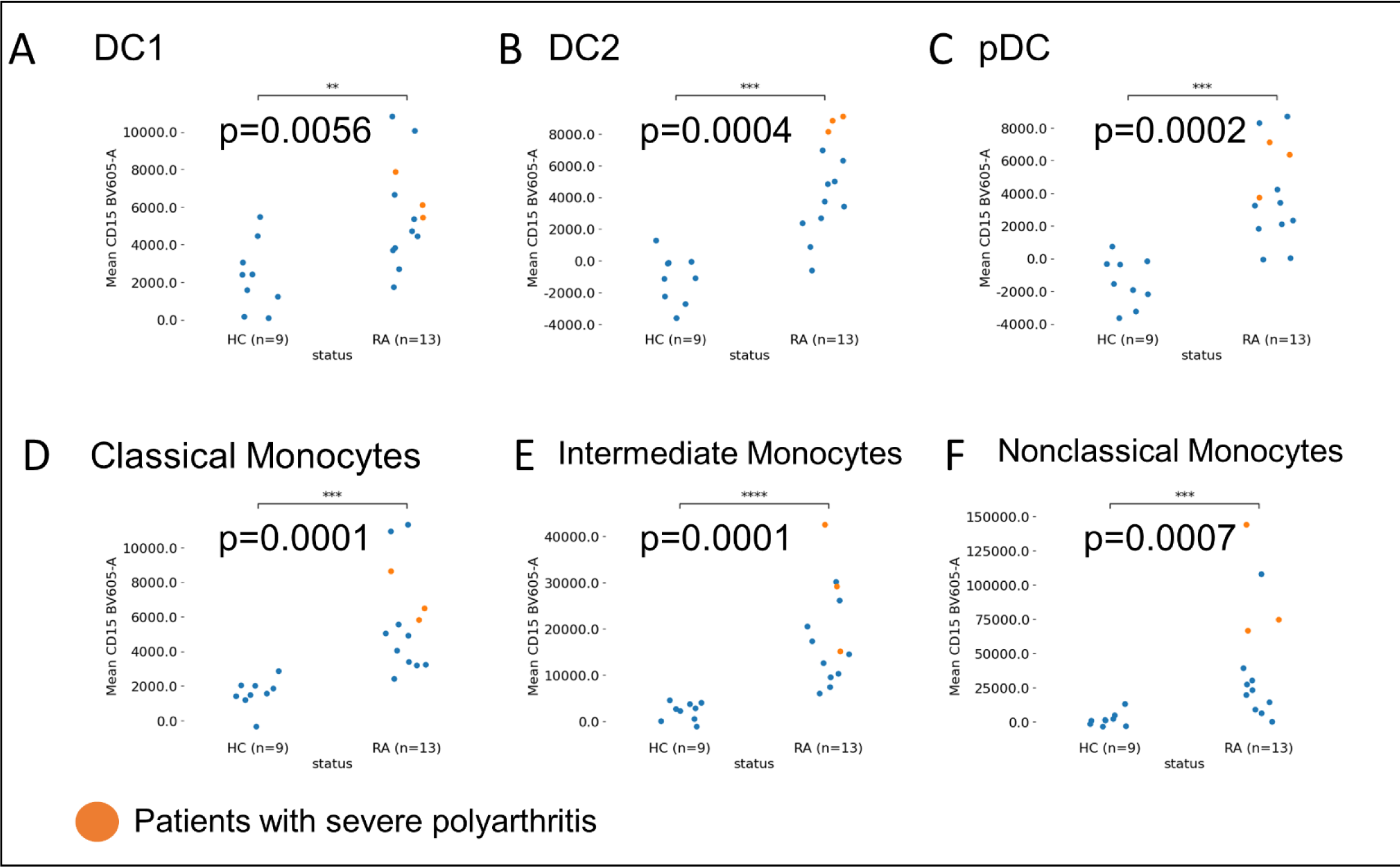
CD15 is increased in antigen-presenting cell (DR^hi^) subsets in RA and healthy controls (HC), in mean fluorescence intensities (MFI) by spectral cytometry. DC1 (CD141^+^) (**A**) DC2 (CD1c^+^) (**B**) Plasmacytoid DC (pDC; CD123^+^CD303^+^) (**C**) Classical monocytes (CD14^hi^CD16^-^) (**D**) Intermediate monocytes (CD14^hi^ CD16^+^) (**E**) Nonclassical monocytes (CD16^+^CD14^lo^) (**F**). RA patients with severe polyarthritis at the time of sample collection are highlighted in orange.

### CD15^+^CD16^+^ phenotypes contribute to the APC (DR^hi^) compartment in RA blood

We adjusted our gating strategy to fully include populations with higher granularity (side scatter, SSC; Supplementary Figure 2). Focusing on CD15, we found that RA patients had a significant expansion of a distinct CD15^+^CD16^+^ population (RA: 1.5% [IQR 0.7 to 3.3], HC: 0.1% [IQR 0.08 to 0.46]; % live leukocytes, p<0.001; Figure 4A, B) compared with healthy control donors. A subset of RA CD15^+^CD16^+^ was HLA-DR positive, contributing significantly to overall DR^hi^ compartment in individuals with RA (CD15^+^CD16^+^DR^hi^, Figure 4B and Supplementary Figure 2G, H).

**Figure 4.**
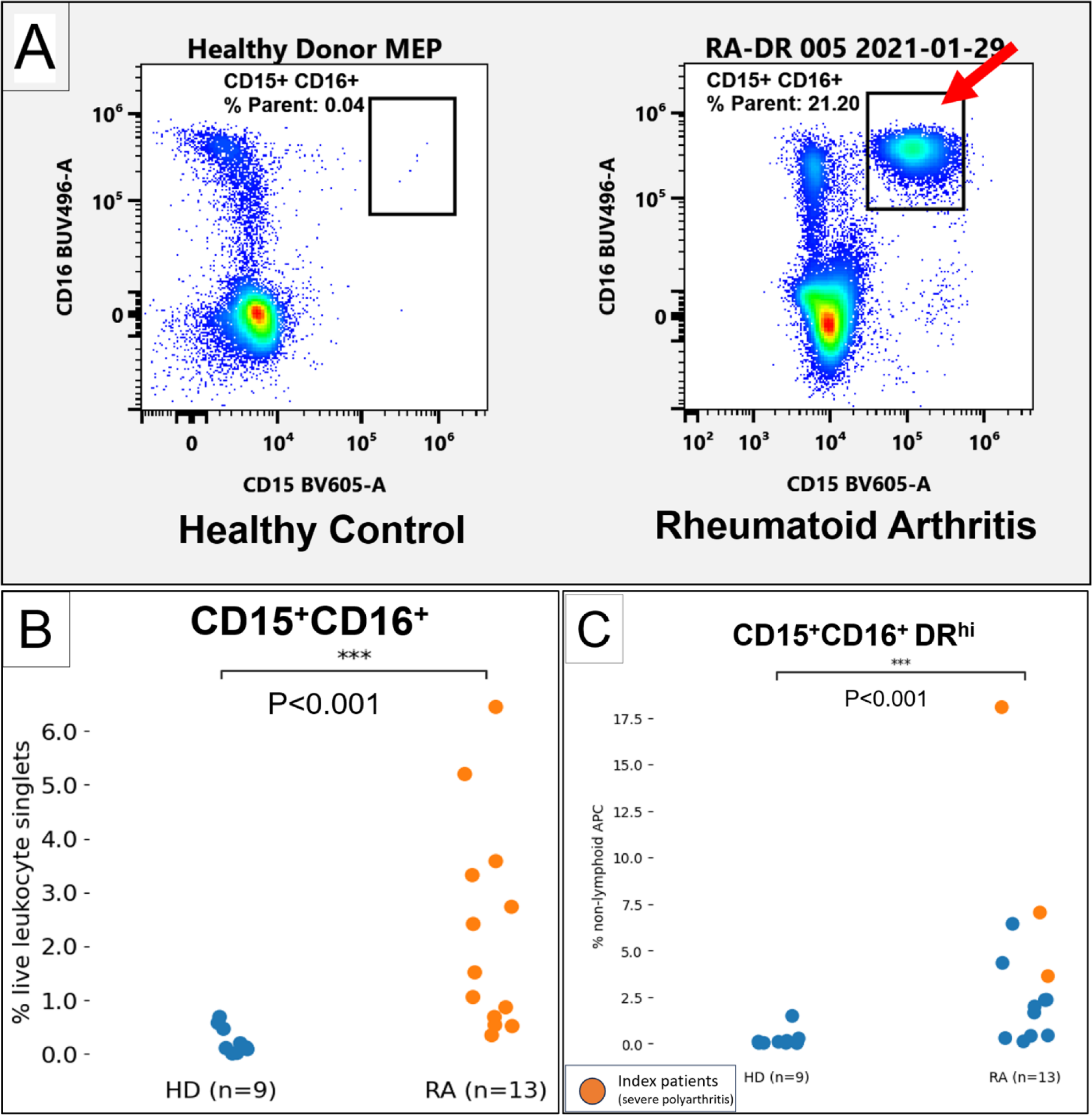
Appearance of a CD15^+^CD16^+^ population in Rheumatoid Arthritis (RA), compared with Health control donors (HD). Representative flow cytometry plots of an active RA patient and a healthy don (HD), gated on live non-lymphoid singlet leukocytes. CD15 (x-axes) and CD16 (y-axes) Left: Health control donor. Right: RA patient with CD15^+^CD16^+^ population (right black gate, red arrow) ( Quantification of CD15^+^CD16^+^, % of live leukocytes in HD (blue) and RA (orange) (**B**) CD15^+^CD16^+^DR cells, in % of non-lymphoid APC ([CD3/19]^-^DR^hi^). RA patients with severe polyarthritis are highlighted

### CD15^+^CD16^+^DR^hi^ exhibit a ‘Hybrid’ phenotype with overlapping granulocytic and monocytic/DC features

We compared the immunophenotype (MFI) of CD15^+^CD16^+^DR^hi^ relative to key markers of the established APC populations in RA (Table 1). CD15, while increased throughout APC populations, far exceeded those of reference APC. Expression of CD14 and CD16 resembled intermediate monocytes but CD15^+^CD16^+^DR^hi^ differed by smaller size (forward scatter, FSC) and higher granularity (side scatter, SSC). CD303 (BDCA-2), generally considered specific for pDC (13), was highly positive, whereas CD123 was much lower, clearly distinguishing these cells from the separate plasmacytoid DC population (Supplementary Figure 10). Given their DR^hi^ phenotype with expression of markers associated with granulocytic/monocytic and plasmacytoid DC lineages we designate them DR^hi^ ‘Hybrid’ cells.

**Table 1.** Phenotype of CD15^+^CD16^+^ DR^hi^ cells (first row) in relationship to reference antigen-presenting cell (DR^hi^) populations. Mean MFI (SD). FSC: Forward Scatter. SSC: Side Scatter.

| Mean MFI<br>(SD) | HLA-DR | CD1c | CD11c | CD14 | CD15 | CD16 | CD19 | CD56 | CD83 | CD86 | CD123 | CCR2 | CCR7 | CD141 | CD163 | CD275 | CD303 | FSC | SSC |
| --- | --- | --- | --- | --- | --- | --- | --- | --- | --- | --- | --- | --- | --- | --- | --- | --- | --- | --- | --- |
| **CD15<sup>+</sup>CD16<sup>+</sup>DR<sup>hi</sup>** | 91.7K<br>(11.5K) | 4223<br>(4581) | 14.1K<br>(7608) | 83.4K<br>(115K) | 157K<br>(83.1K) | 169K<br>(73.3K) | -218<br>(3113) | 5925<br>(434) | 15.5K<br>(12.6K) | 3559<br>(2393) | 11.2K<br>(17.3K) | 165K<br>(109K) | 4896<br>(2332) | 18.5K<br>(12.4K) | 9639<br>(3130) | 76.7K<br>(45.6K) | 179K<br>(67.9K) | 1.38M<br>(410K) | 1.61M<br>(240K) |
| **B cells** (CD19<sup>+</sup>) | 79.9K<br>(29.6K) | 10.2K<br>(4793) | -1631<br>(1706) | 938<br>(920) | 2696<br>(2157) | -3131<br>(2351) | 17.3K<br>(6005) | 1598<br>(377) | -1984<br>(3174) | 2252<br>(655) | 7470<br>(4928) | 3736<br>(1647) | 9760<br>(2125) | 4998<br>(1183) | 1981<br>(484) | 3549<br>(1743) | 2810<br>(3087) | 996K<br>(83.7K) | 342K<br>(27.0K) |
| **DC1** (CD141<sup>+</sup>) | 171K<br>(46.6K) | 3031<br>(1926) | 3548<br>(4840) | 987<br>(13.8K) | 4813<br>(2885) | -3932<br>(5845) | -1097<br>(477) | 10.6K<br>(8863) | 1586<br>(2411) | 3453<br>(1227) | 17.0K<br>(16.0K) | 55.4K<br>(18.7K) | 3776<br>(1908) | 587K<br>(112K) | 1672<br>(796) | 4796<br>(2056) | 2624<br>(1790) | 1.89M<br>(224K) | 1.25M<br>(161K) |
| **DC2** (CD1c<sup>+</sup>) | 229K<br>(30.0K) | 62.8K<br>(24.2K) | 23.9K<br>(18.2K) | 10.5K<br>(3316) | 4722<br>(3134) | -1030<br>(3248) | -1179<br>(1201) | 9150<br>(8124) | -36<br>(3838) | 5579<br>(1708) | 28.4K<br>(12.0K) | 220K<br>(39.1K) | 3535<br>(1102) | 45.9K<br>(8998) | 6668<br>(1533) | 3700<br>(2016) | 1778<br>(2729) | 1.86M<br>(177K) | 1.02M<br>(141K) |
| **pDC**<br>(CD303<sup>+</sup> CD123<sup>+</sup>) | 125K<br>(25.3K) | 7047<br>(4583) | -1596<br>(2799) | 5140<br>(3823) | 717<br>(1613) | 160<br>(2447) | -2991<br>(2008) | 6726<br>(3602) | 1368<br>(2677) | 3438<br>(1312) | 66.3K<br>(19.5K) | 182K<br>(50.9K) | 4348<br>(1062) | 52.9K<br>(17.5K) | 6023<br>(1524) | 2794<br>(1363) | 50.8K<br>(11.5K) | 1.91M<br>(152K) | 910K<br>(98.3K) |
| **Classical monocytes**<br>(CD14<sup>hi</sup> CD16<sup>-</sup>) | 127K<br>(10.8K) | 6124<br>(2879) | 21.5K<br>(14.8K) | 251K<br>(47.9K) | 6073<br>(2500) | 2000<br>(5590) | -2547<br>(1557) | 5261<br>(5757) | 71<br>(6433) | 14.4K<br>(4351) | 9267<br>(15.5K) | 346K<br>(58.3K) | 3214<br>(1816) | 35.3K<br>(11.5K) | 7586<br>(1727) | 8555<br>(3785) | 109<br>(1737) | 2.00M<br>(157K) | 1.17M<br>(195K) |
| **Non-classical monocytes**<br>(CD14<sup>lo</sup> CD16<sup>+</sup>) | 122K<br>(24.5K) | 10.3K<br>(7897) | 45.5K<br>(27.1K) | 16.8K<br>(4042) | 42.6K<br>(44.0K) | 132K<br>(50.7K) | -610<br>(884) | 5239<br>(3806) | 345<br>(6830) | 15.9K<br>(6822) | 24.6K<br>(13.9K) | 51.7K<br>(37.4K) | 5744<br>(2251) | 31.1K<br>(11.4K) | 5300<br>(1986) | 18.0K<br>(25.4K) | 31.8K<br>(39.6K) | 1.81M<br>(235K) | 1.13M<br>(141K) |
| **Intermediate monocytes**<br>(CD14<sup>hi</sup> CD16<sup>+</sup>) | 143K<br>(34.2K) | 6595<br>(3022) | 36.5K<br>(21.3K) | 196K<br>(65.8K) | 18.4K<br>(10.8K) | 49.8K<br>(13.7K) | -2208<br>(1386) | 6065<br>(6091) | -339<br>(7174) | 21.0 K<br>(8231) | 16.2K<br>(16.6K) | 239K<br>(109K) | 5449<br>(4545) | 42.7K<br>(8988) | 8237<br>(5387) | 9307<br>(4360) | 6455<br>(7633) | 2.09M<br>(159K) | 1.34M<br>(187K) |

### CD15^+^CD16^+^DR^hi^ ‘Hybrid’ cells are enriched in the blood of RA patients with severe polyarthritis

We then focused on these 3 highlighted RA patients with early, active, debilitating RA polyarthritis (Figure 4C, orange and Supplementary Table 6). t-SNE analyses of CD15^+^ CD16^+^ of these HLA-DR highlight co-expression of CD303 and CD83 while affected by severe polyarthritis (Figure 5). Flow cytometry after effective therapy showed resolution of most CD15^+^CD16^+^ with only few residual DR^hi^ events with a resulting scattered pattern (Supplementary Figure 11).

**Figure 5.**
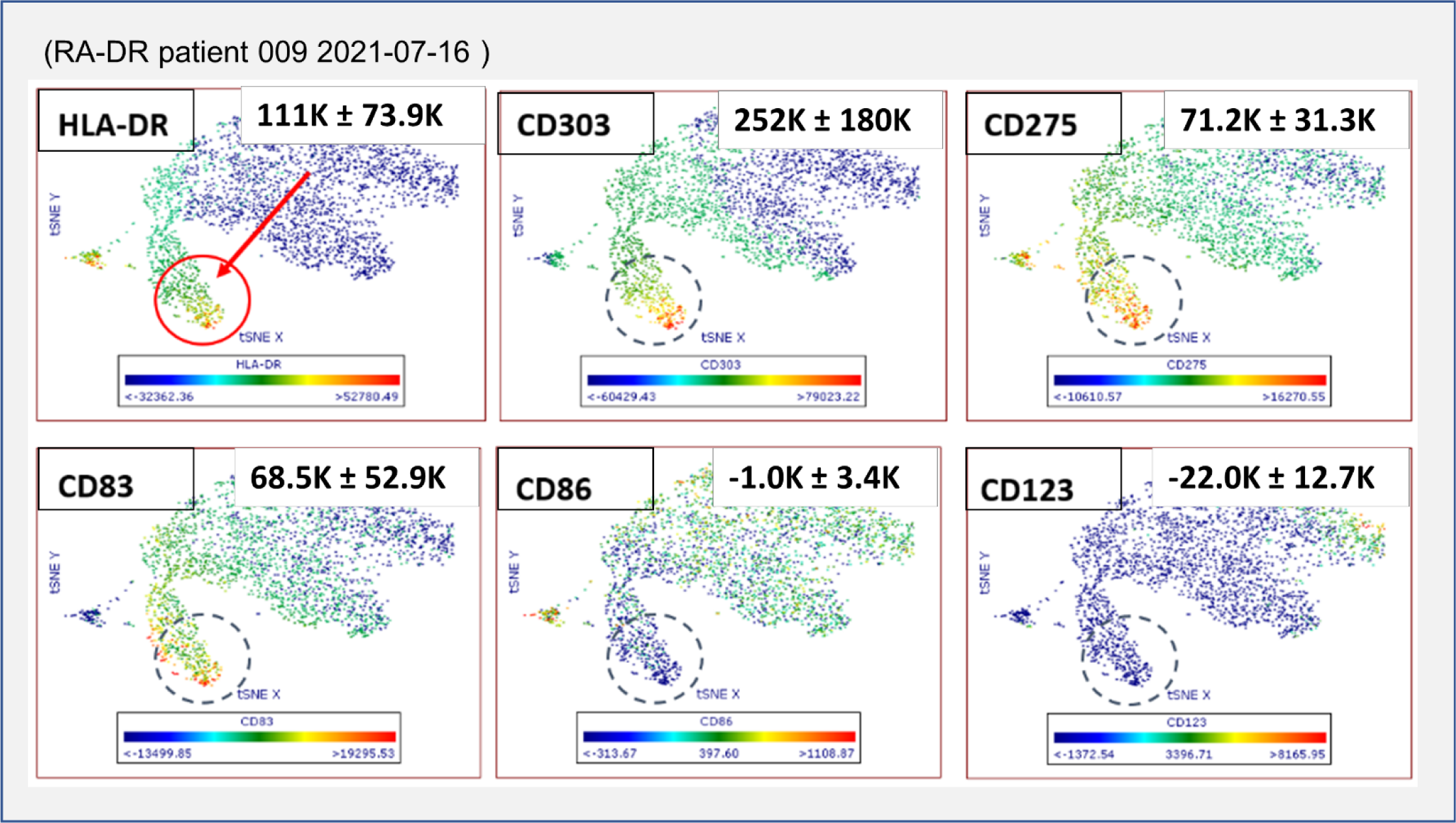
t-SNE analysis of CD15^+^CD16^+^ cells. CD15^+^CD16^+^ cells include an HLA-DR positive cluster (red circle). Cells were CD303 (BDCA-2) positive while low for CD123. CD275 (ICOS-L) and CD83 were positive; CD86 was not expressed (dashed circlesColors represent mean fluorescence intensities (MFI). Blue: low MFI. Red: High MFI. Mean MFI of the circled population displayed in right upper boxes. ). Sample from patient with polyarthritis at time of collection, a representative t-SNE plot from the RA patient after effective therapy is shown in Supplementary Figure 11.

## DISCUSSION

Here, we used HLA-DR to capture APC for spectral cytometry. This comprehensive perspective documents widespread alterations of the APC landscape in RA, particularly for CD1c^+^ dendritic cells. The striking association of a shared HLA-DR (MHC II) sequence with RA, and the fact that MHC II molecules are involved in the capture and subsequent processing of antigens, point towards a central role of DR^hi^ cells in RA. We had thus anticipated increased HLA-DR expression within CD1c^+^ dendritic cells and other DR^hi^ populations. However, our findings show a different picture.

Except for B lymphocytes (where HLA-DR was increased, as anticipated) HLA-DR did not differ between RA and healthy control APC, arguing against an overall increase in antigen-presentation in RA. And, while we had expected expanded CD1c^+^ dendritic cells, we found that the contribution of CD1c^+^ dendritic cells to the overall RA APC (DR^hi^) compartment is in fact decreased. Instead, we show that alterations in CD1c^+^ dendritic cells (DC2) include increased co-stimulatory molecules CD86, CD275 (ICOS- L) and chemokine receptor CCR7. Our findings also suggest that CD15^+^CD16^+^ DR^hi^ cells contribute to the APC compartment in RA, providing a more comprehensive picture of changes in RA APC. One possibility is that they represent low density granulocytes, described in Lupus (14, 15) and other inflammatory diseases (16) but, given their phenotypic features overlap with monocytes and dendritic cells, they are similarly reminiscent of a neutrophil-DC ‘hybrid’ cell reported in multiple sclerosis (17). Their co-stimulatory phenotype suggests these cells can participate in altered co-stimulatory signals to CD4^+^ T cells and thus participate in adaptive immunity by providing proinflammatory effects on T cells (Figure 6). This model reconciles the otherwise paradoxical observation of decreased dendritic cell subsets in RA (18).

**Figure 6.**
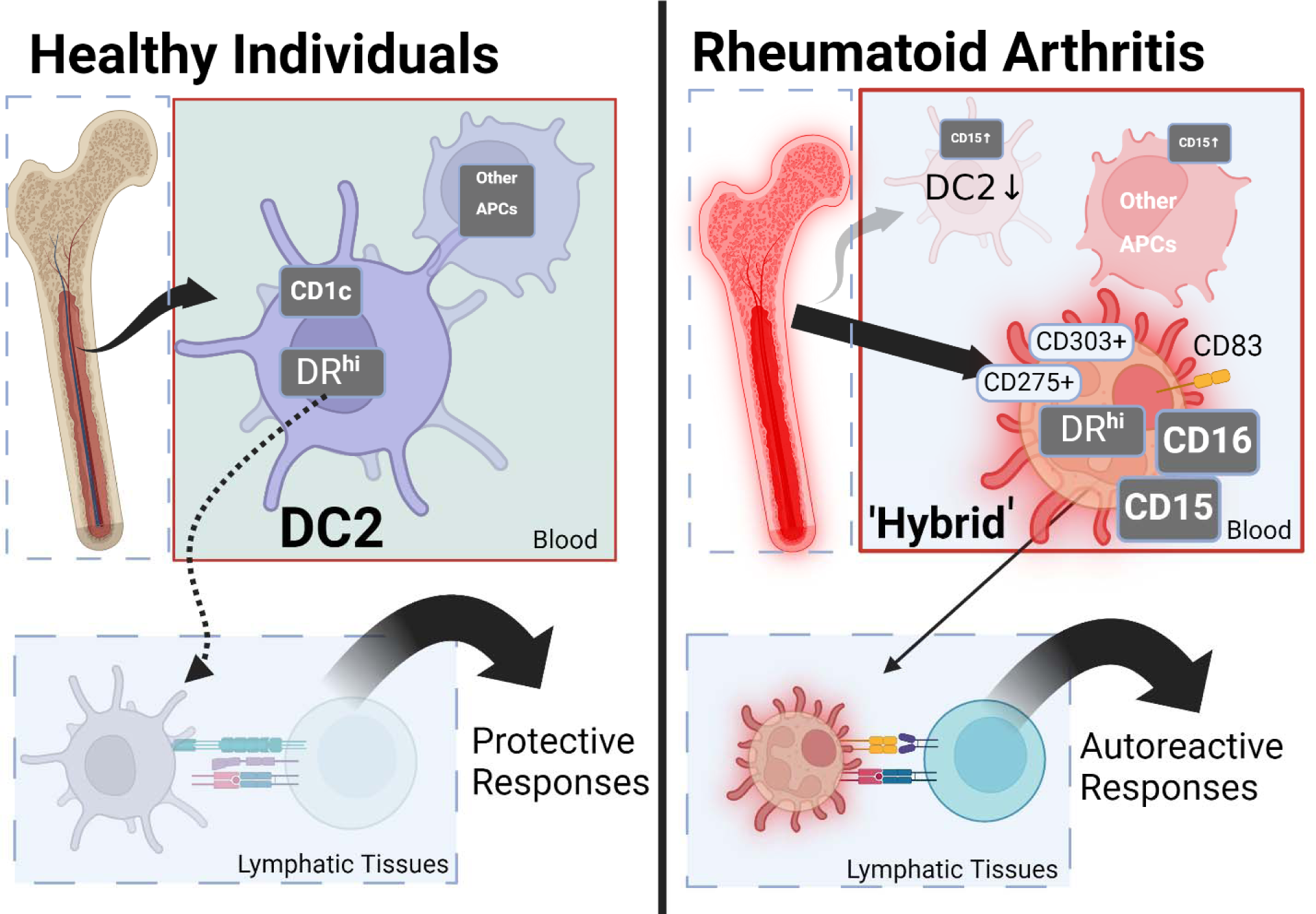
A model of how changes in DR^hi^ compartment and emergence of DR^hi^ CD15 CD16 cells can contribute to the pathophysiology of rheumatoid arthritis. Made using Biorender. Left: In healthy individuals the blood APC (DR^hi^) compartment consists of DC2 and other ‘established’ APC populations, providing protective immunity. Right: The APC (DR^hi^) compartment in RA is characterized by a decrease and inflammatory phenotype of DC2. Additionally, there is an appearance of CD15^+^CD16^+^ DR^hi^ ‘Hybrid’ cells with a co-stimulatory phenotype, suggesting involvement of these DR^hi^ cells in altered antigen presentation to T lymphocytes.

Strengths of our study include a comprehensive, biologically plausible, perspective that retains all HLA-DR expressing cells for analysis. It adds new facets by bringing the attention to apparently new DR^hi^ phenotypes including those with granulocytic features. Limitations include sample size and the focus on blood; assessing the functional properties of the co-stimulatory CD15^+^ phenotype will be an important future aspect. Yet our current findings highlight how shifts in RA blood APC with co-stimulatory potential are not restricted to lymphocytes, monocytes, and dendritic cells.

These and other ‘unorthodox’ DR^hi^ phenotypes could explain aspects of autoimmune diseases and represent potential new research and therapeutic targets. The granular side scatter profile and CD14, CD15, CD16 phenotype of these cells hints they are of granulo-monocytic bone marrow origin whereas increased CCR7 in DC suggests a lymphoid tissue destination.We hope our study will encourage a fresh and inclusive look on existing and emerging data including and beyond the described CD15^+^CD16^+^DR^hi^ cells in blood and other tissues.

## Author contributions

CG devised, performed and analyzed cytometry experiments, recruited patients and controls, and wrote the manuscript; he is the guarantor. HQ performed cytometry experiments and t-SNE analyses. JBG screened and recruited patients. RW gave essential methodological and conceptual guidance. AP contributed to the design of the study, data analyses, and edited the manuscript. All authors contributed to the study design.

## Data availability

The data relevant to the study are included in the article or are available in the online supplement. The raw spectral cytometry files are available upon reasonable request.

## Ethics approval and patient consent

Approved by the institutional review board of SUNY Upstate Medical University (Syracuse, USA), no. 1659139. All patients and healthy volunteers consented to participate in the study.

## Permission to reproduce material from other sources

Not applicable

## Clinical trial registration

Not applicable

## Acknowledgments

We thank the Flow Cytometry Core of SUNY Upstate Medical University for providing equipment support and the patient and healthy volunteers for their participation.

