## Supplementary materials and methods for "CD1c Dendritic Cells are depleted and accompanied by new HLA-DR^hi^ Phenotypes in Rheumatoid Arthritis Blood"

### Supplementary appendix

Gating strategy

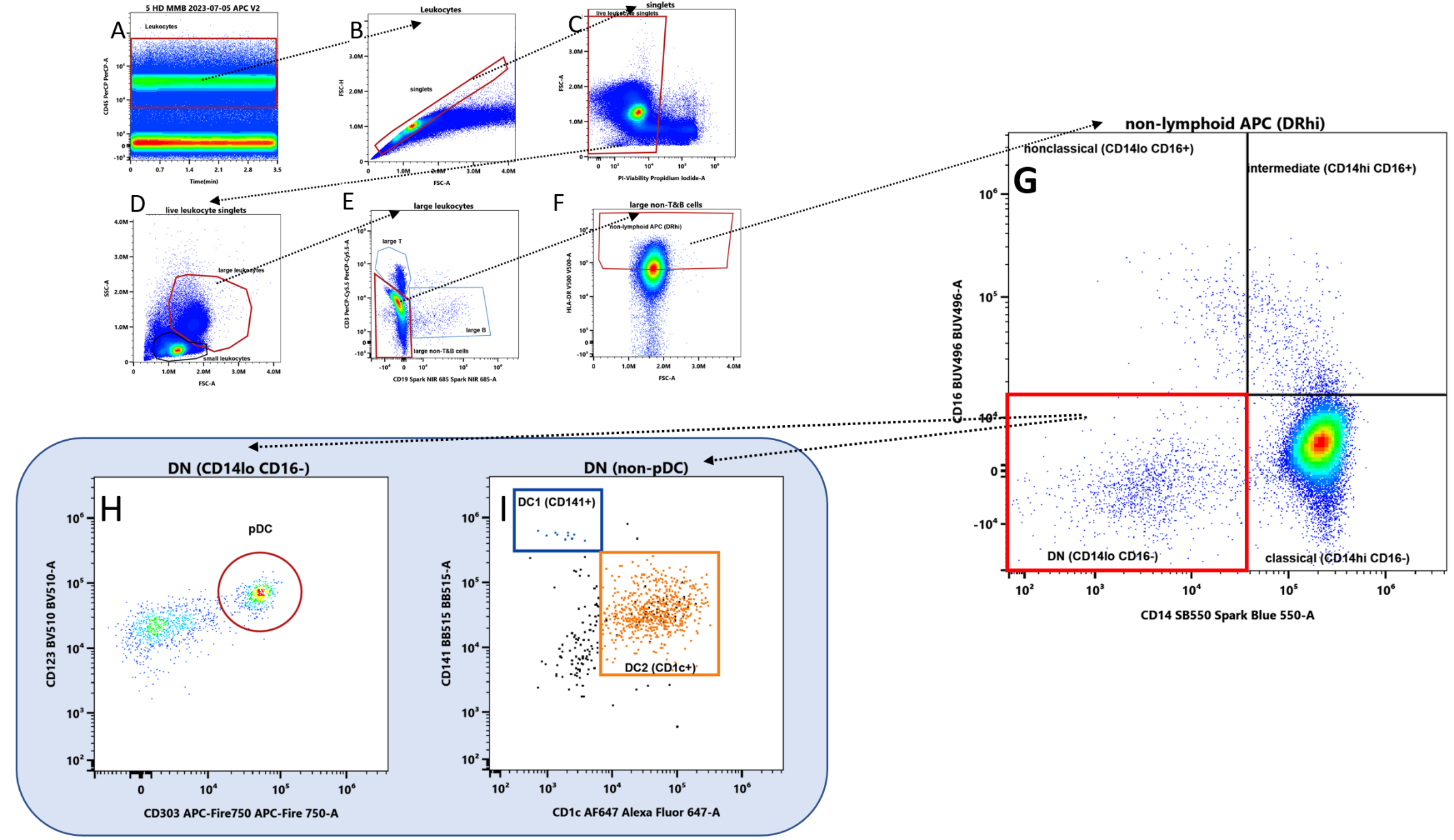

**Supplementary Figure 1** Gating Strategy for non-lymphoid reference APC (Monocytes, plasmacytoid DC, DC1 (CD141+) and DC2 (CD1c+)). We gated large live singlet CD45+ leukocytes (A-D). Lymphoid cells (CD3+, CD19+) and HLA-DRneg/lo cells were gated out (E, F). Classical, intermediate, and non-classical monocytes were gated by CD14 and CD16 expression (G). We then gated pDC as CD303+CD123+ population (H). After applying a NOT(pDC) gate (that removes pDC gated in H), we gated CD141+ DC1 (blue events) and CD1c+ DC2 (orange events) (I). Healthy donor shown.

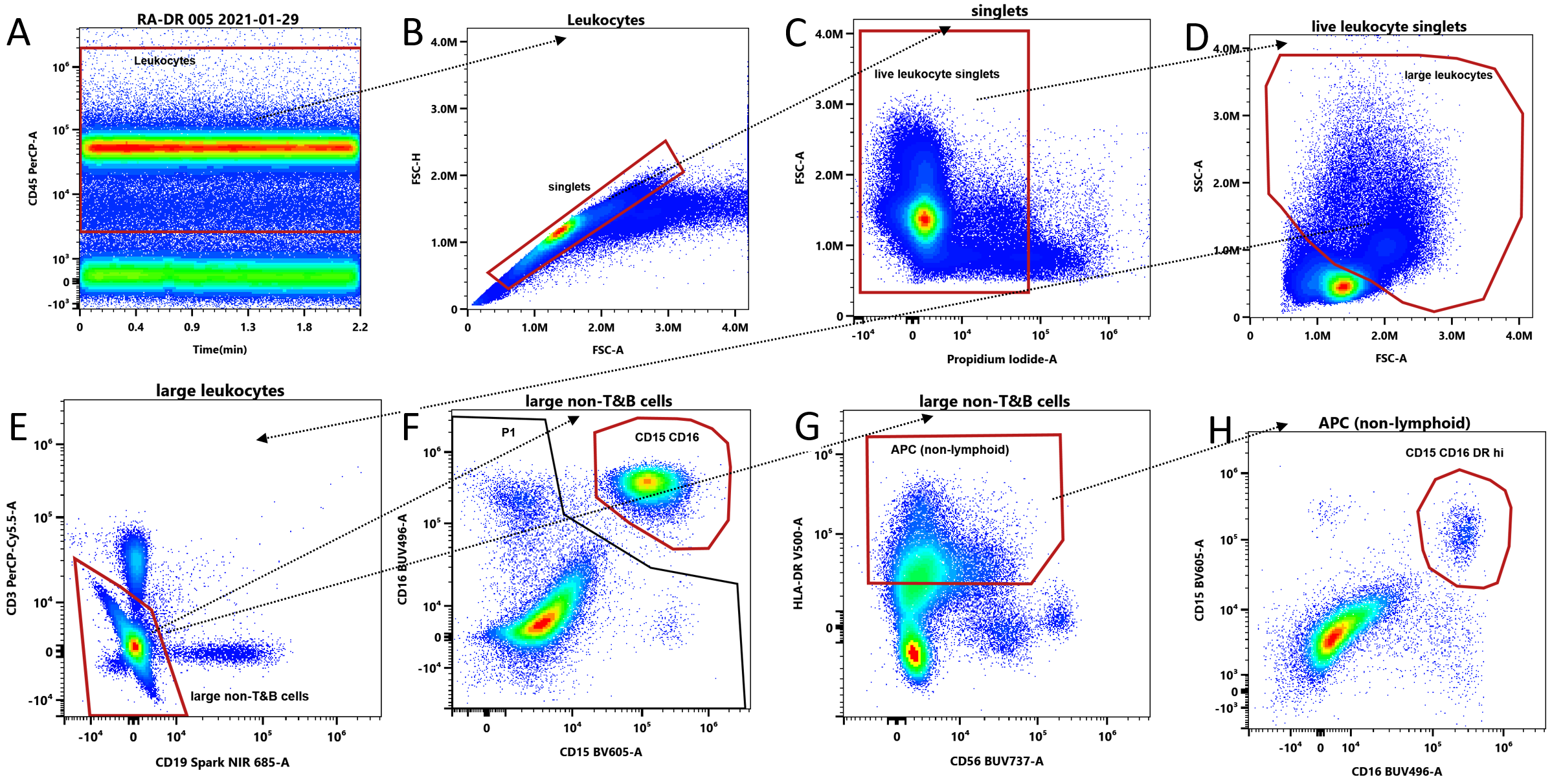

**Supplementary Figure 2** Gating Strategy for CD15^+^CD16^+^ and CD15+CD16+DR^hi^. We gated live singlet CD45+ leukocytes (A-C). The “large leukocytes” gate was widened to ensure full capture of granulocytic populations (D, red gate and dashed ellipse). Lymphoid cells (CD3+, CD19+) were gated out (E). CD15+CD16+ cells were gated (F). Separately, large non-T&B cells shown in E were gated for HLA-DR^hi^, HLA-DRneg/lo cells were gated out (G). The CD15+CD16+ DR^hi^ population was gated (H). (E, F). RA patient with polyarticular synovitis shown.

A

B

C

D

E

F

G

H

I

J

K

L

M

N

O

P

Q

R

S

T

U

V

W

X

Y

**Supplementary Figure 3** CD141^+^ Dendritic Cells (DC1) — Phenotypic comparison between RA patients (blue) and healthy donors (orange).

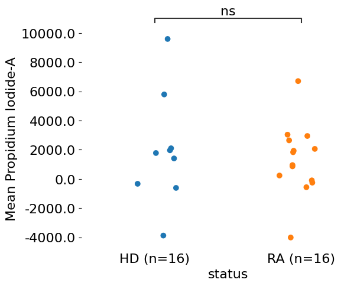

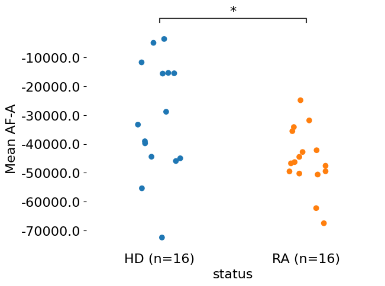

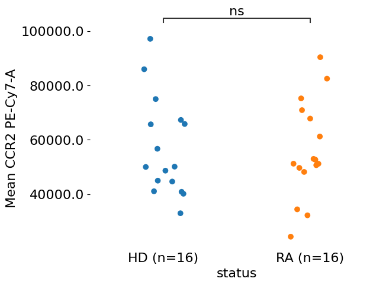

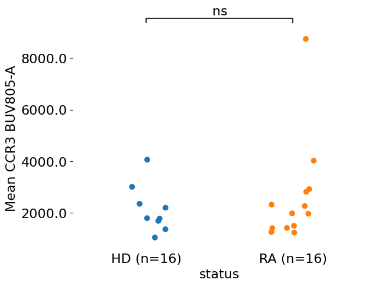

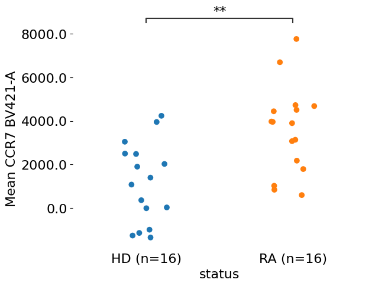

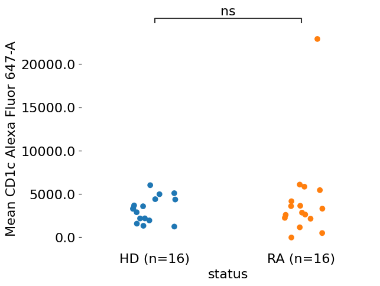

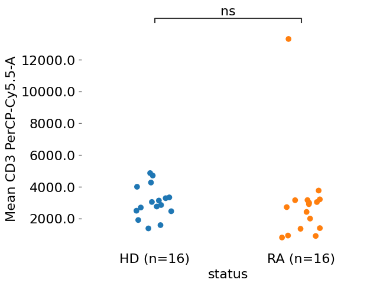

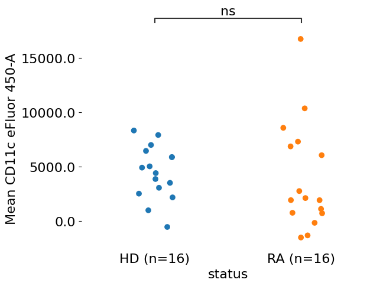

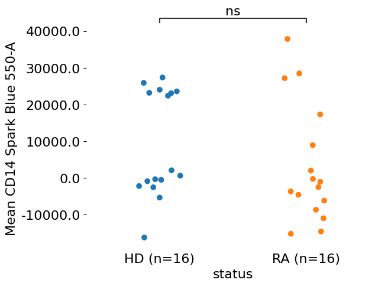

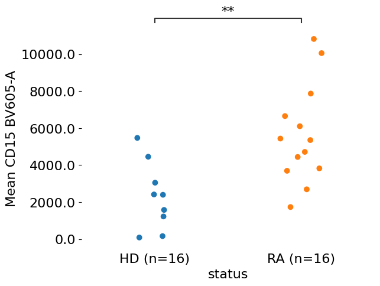

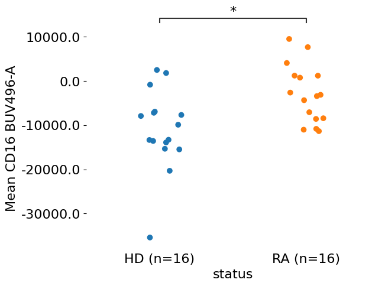

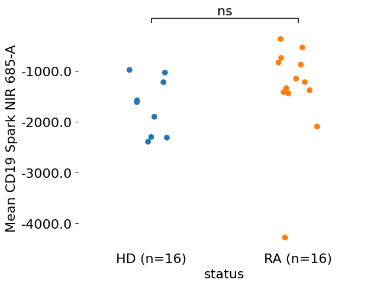

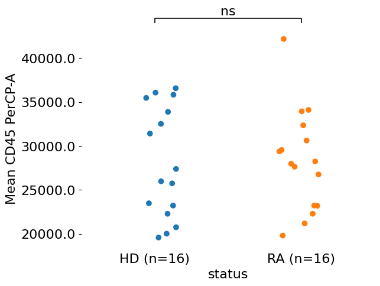

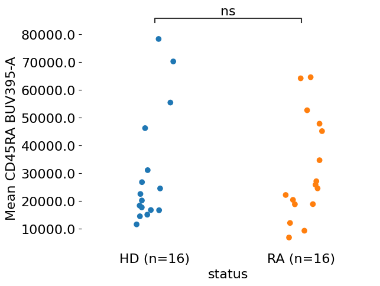

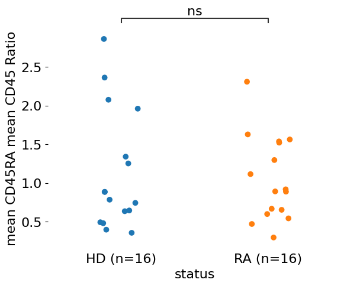

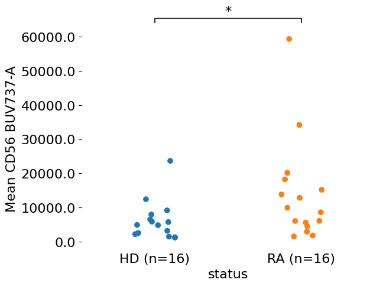

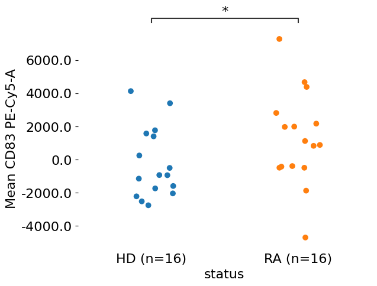

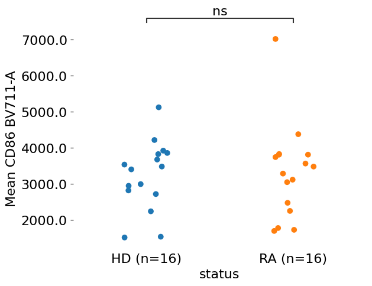

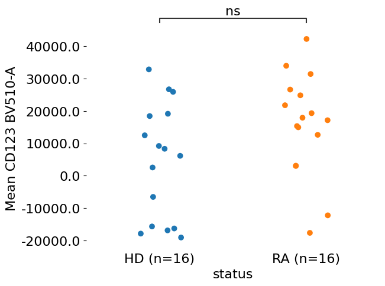

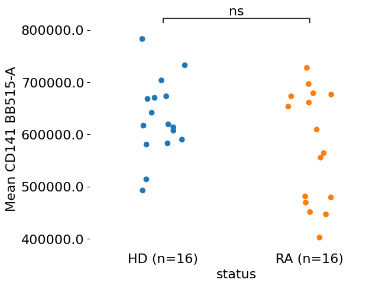

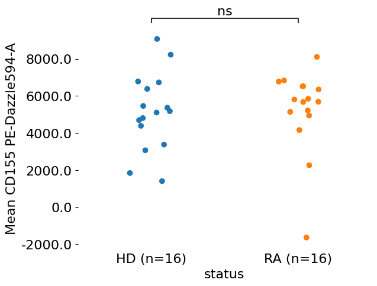

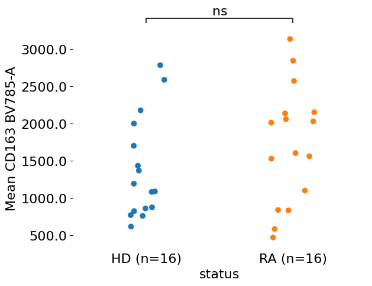

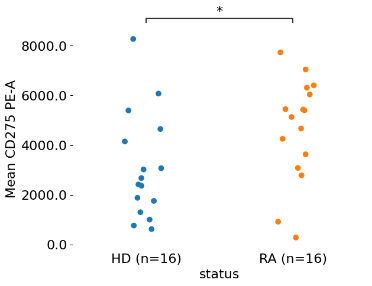

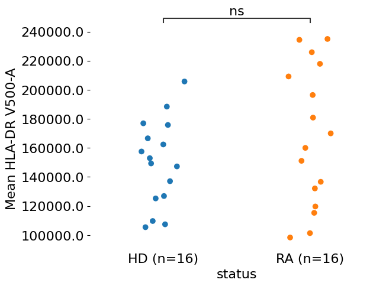

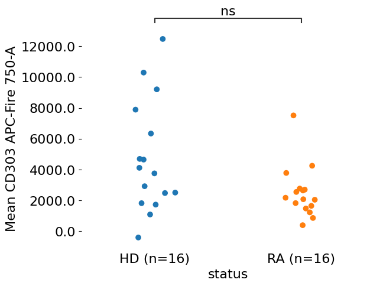

Y

X

W

V

U

T

S

R

Q

P

O

N

M

L

K

J

I

H

G

F

E

D

C

B

A

**Supplementary Figure 4** CD1c^+^ Dendritic Cells (DC2) — Phenotypic comparison between RA patients (blue) and healthy donors (orange).

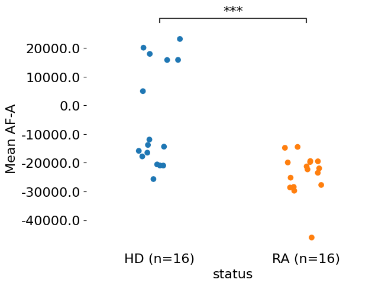

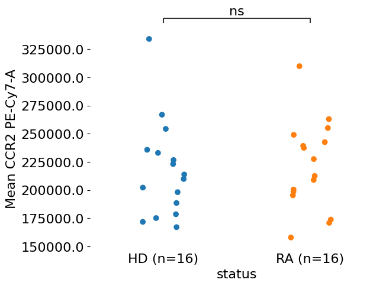

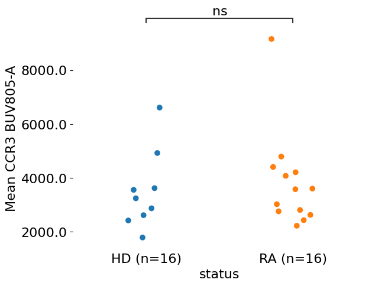

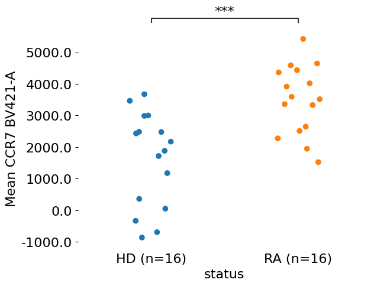

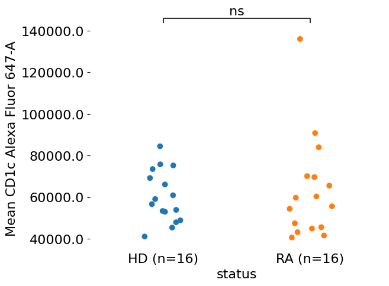

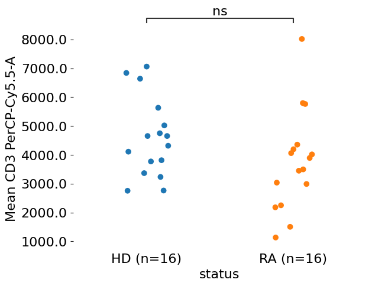

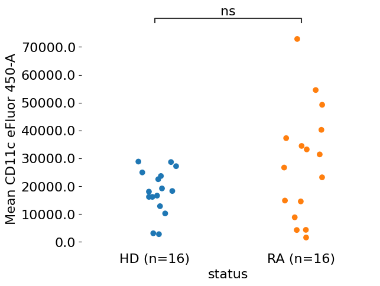

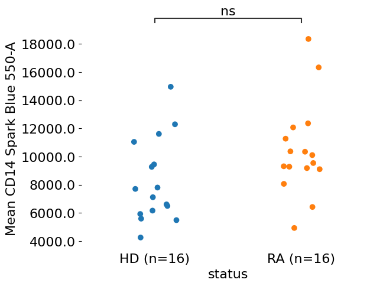

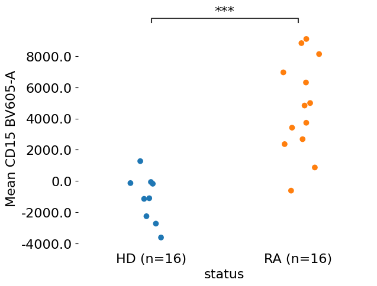

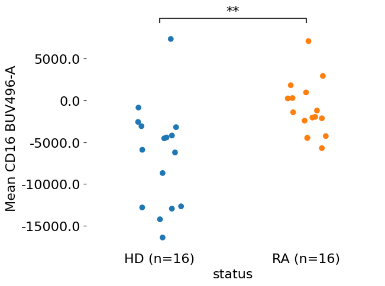

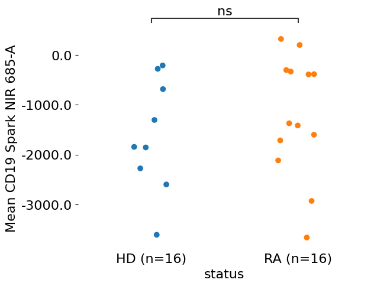

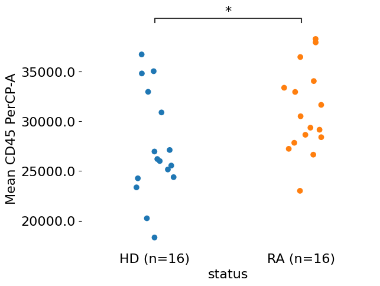

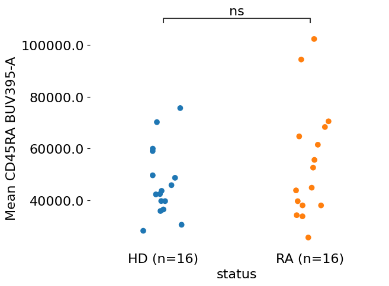

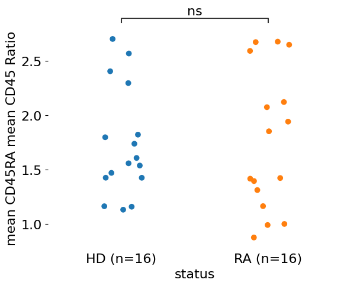

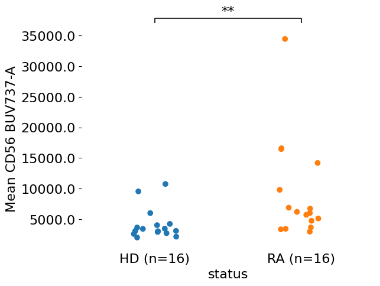

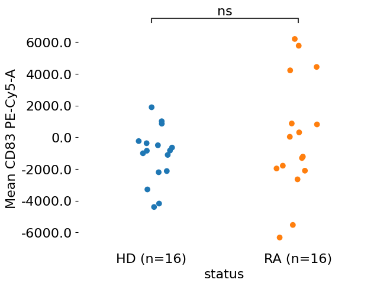

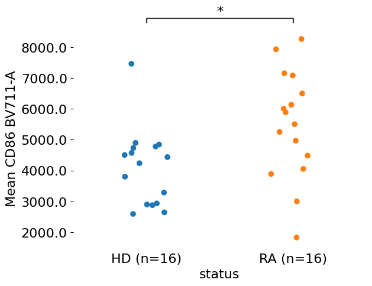

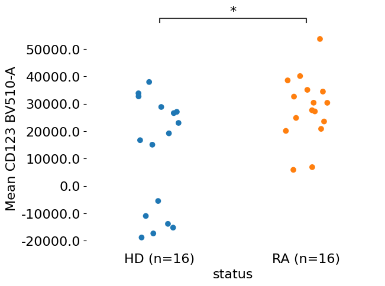

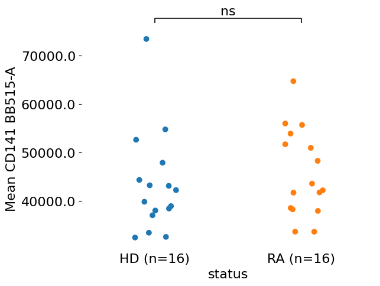

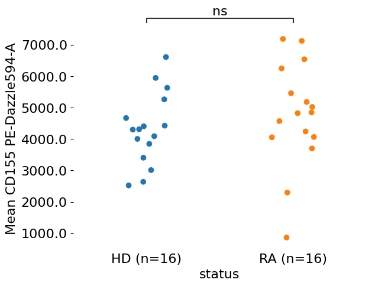

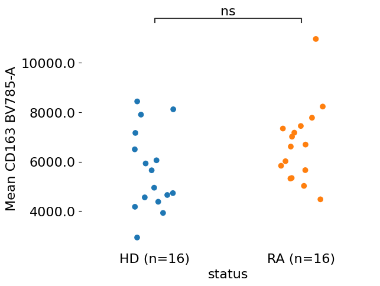

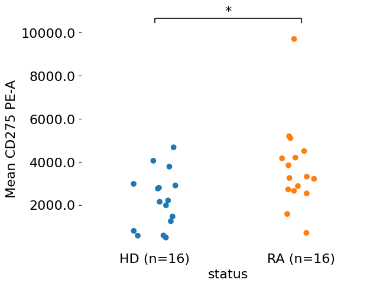

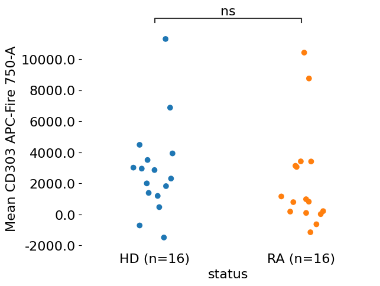

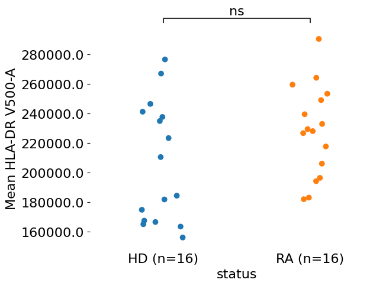

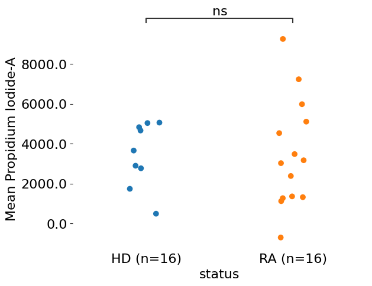

**Supplementary Figure 5**. Plasmacytoid Dendritic Cells (pDC) — Phenotypic comparison between RA patients (blue) and healthy donors (orange).

A

B

C

D

E

F

G

H

I

J

K

L

M

N

O

P

Q

R

S

T

U

V

W

X

Y

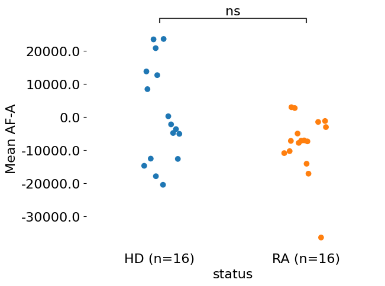

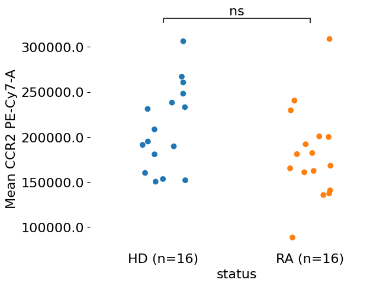

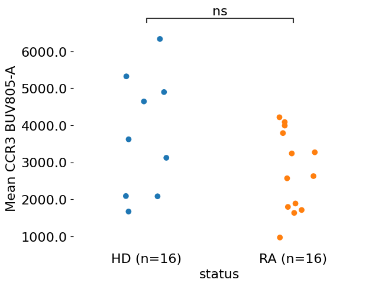

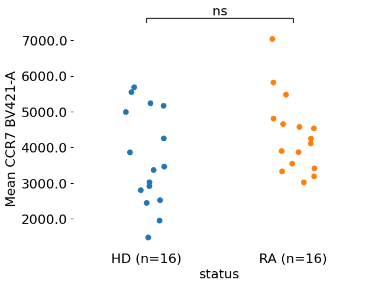

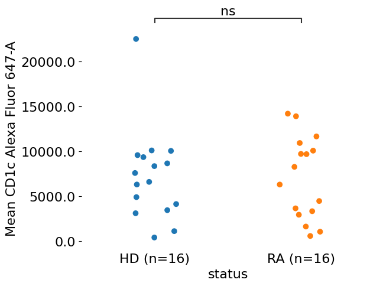

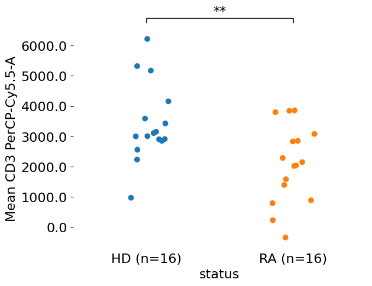

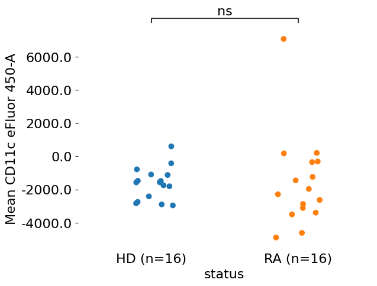

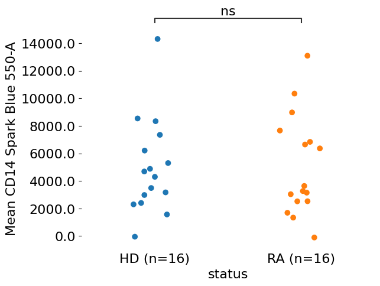

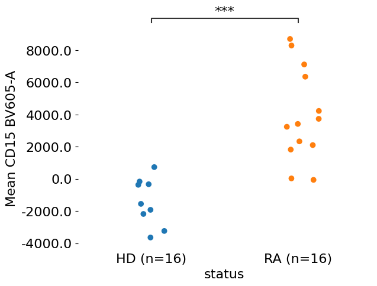

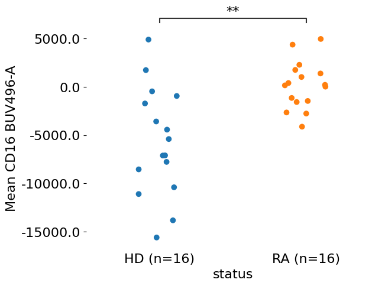

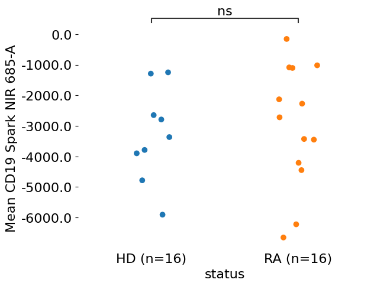

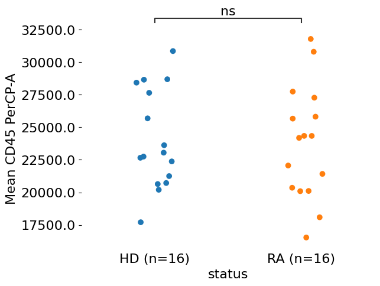

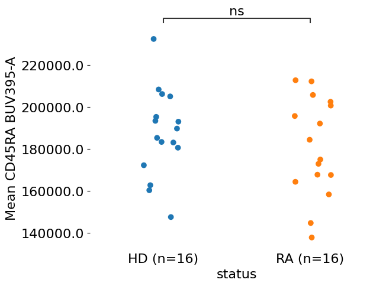

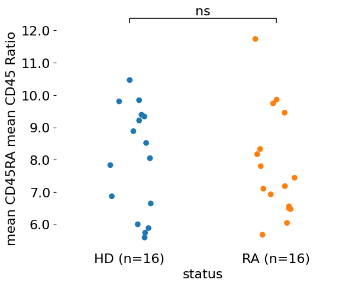

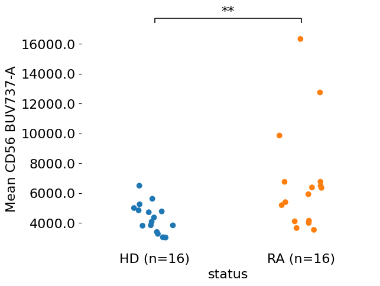

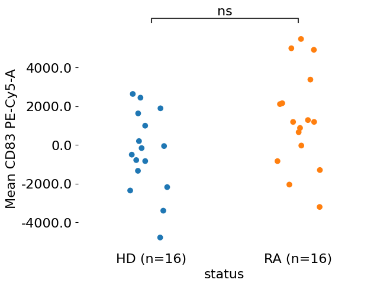

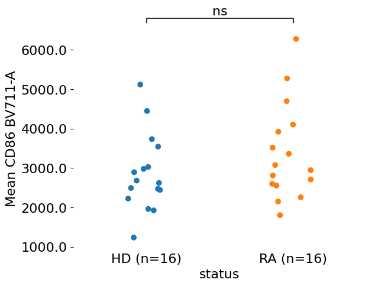

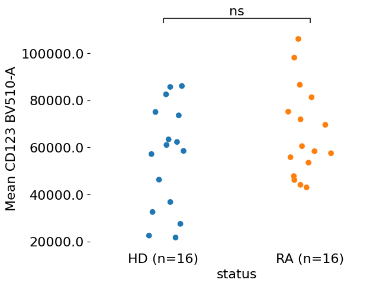

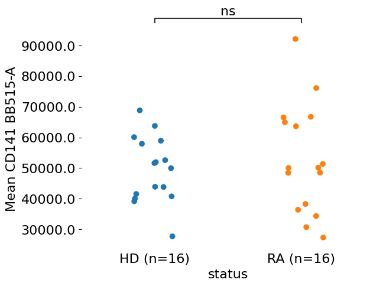

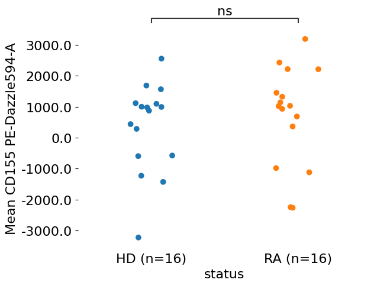

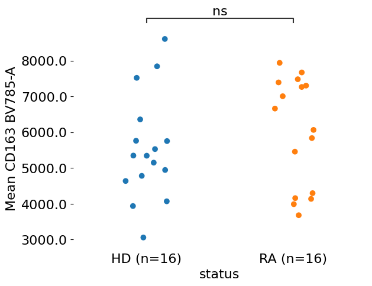

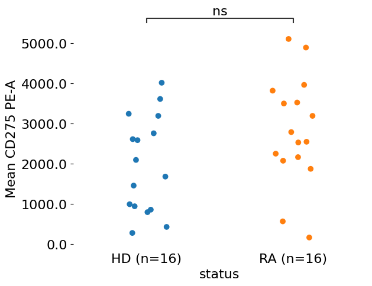

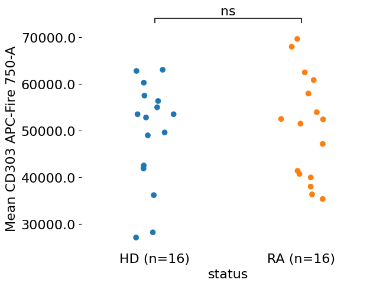

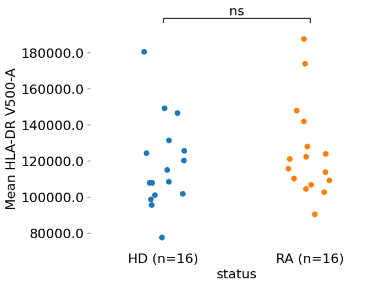

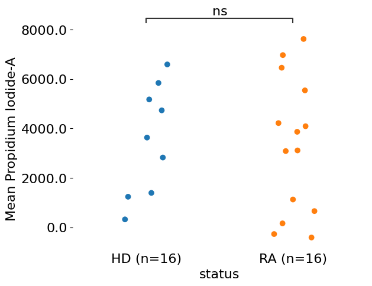

**Supplementary Figure 6** CD19^+^ B cells — Phenotypic comparison between RA patients (blue) and healthy donors (orange).

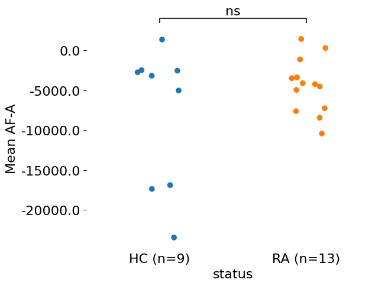

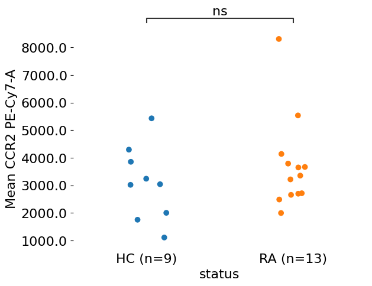

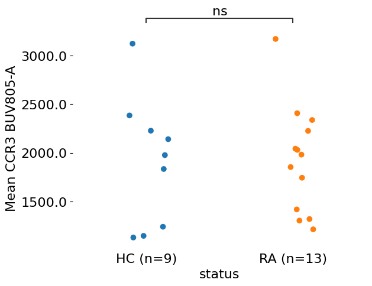

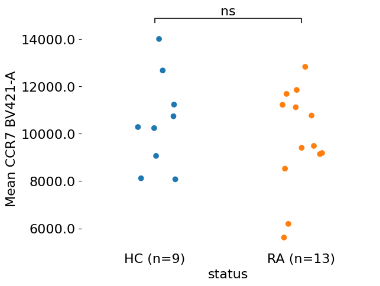

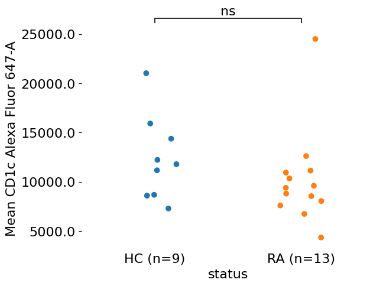

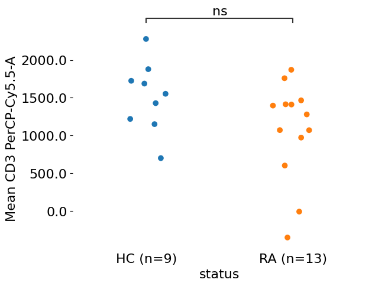

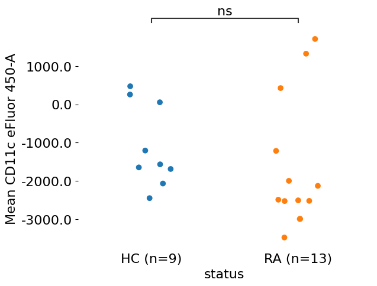

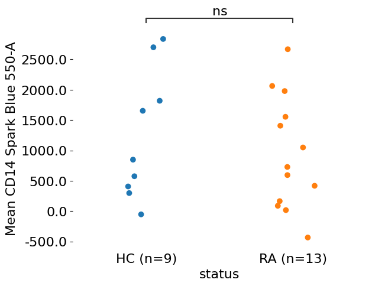

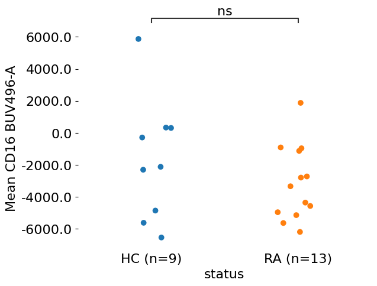

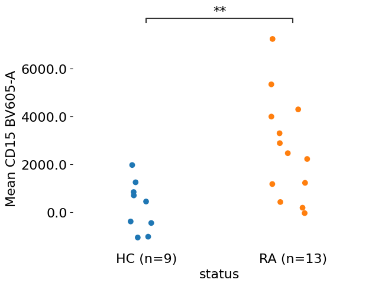

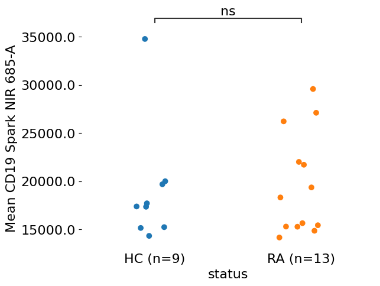

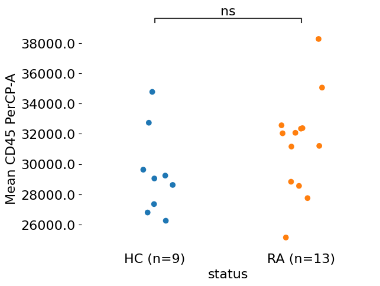

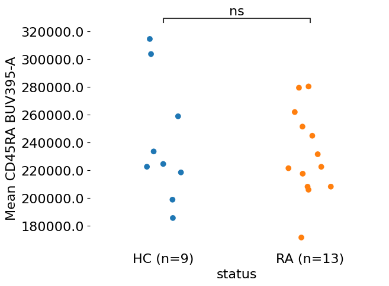

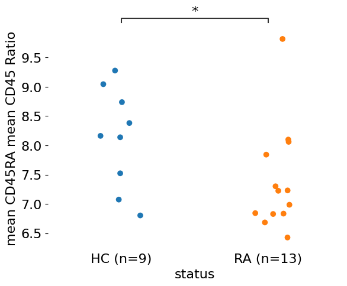

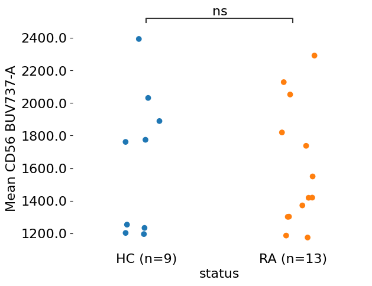

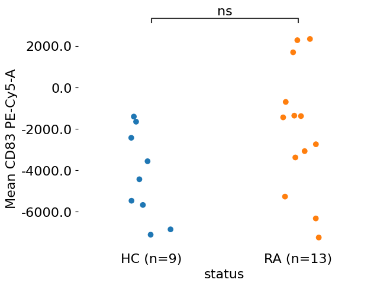

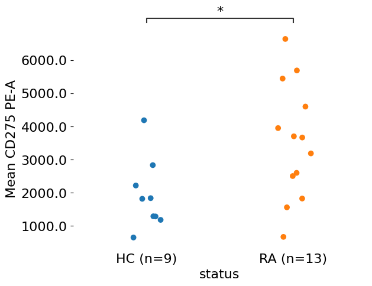

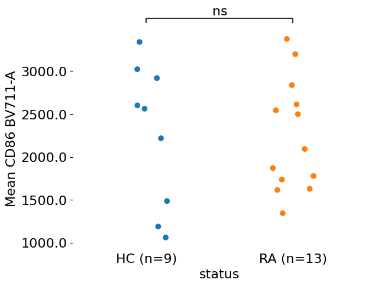

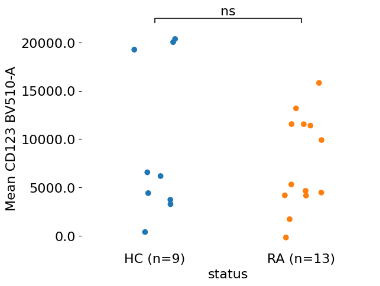

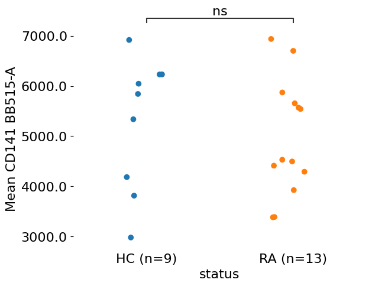

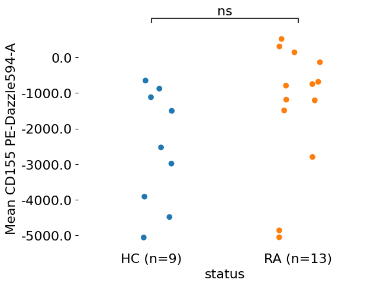

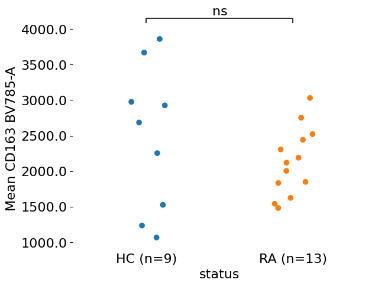

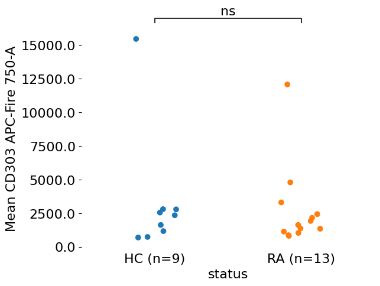

A

B

C

D

E

F

G

H

I

J

K

L

M

N

O

P

Q

R

S

T

U

V

W

X

Y

A

B

C

D

E

F

G

H

I

J

K

L

M

N

O

P

Q

R

S

T

U

V

W

X

Y

**Supplementary Figure 7** Classical Monocytes (CD14^+^CD16^-/lo^)

A

B

C

D

E

F

G

H

I

J

K

L

M

N

O

P

Q

R

S

T

U

V

W

X

Y

**Supplementary Figure 8** Intermediate monocytes (CD14^+^CD16^+^)

A

B

C

D

E

F

G

H

I

J

K

L

M

N

O

P

Q

R

S

T

U

V

W

X

Y

**Supplementary Figure 9** Nonclassical monocytes (CD14^-^CD16^+^)

**Supplementary Figure 10**. Representative flow cytometry gated on DR^hi^ (CD3^-^CD19^-^) cells in RA and healthy donor.
Gated are plasmacytoid DC (CD123^+^CD303^+^, black gate) and DR^hi^CD303^+^CD123^-^ (red gate).
SSC: Side scatter. RA: rheumatoid arthritis. HD: Healthy control donor.

**Supplementary Figure 11** Representative t-SNE of CD15 CD16 DRhi after effective RA therapy.

**Supplementary Tables**

|  | RA (n=16) | Healthy Controls (n=16) |
| --- | --- | --- |
| Age (mean, SD) | 54.6 ± 16.2 years | 54.0 ± 10.6 years |
| Gender (%, n) |  |  |
| Female | 75% (12) | 81.3% (13) |
| Male | 25% (4) | 18.8% (3) |
| Other | 0% (0) | 0% (0) |
| Ethnicity (%, n) |  |  |
| White | 75% (12) | 93.8% (15) |
| Black | 12.5% (2) | 0% (0) |
| Native American | 12.5% (2) | 0% (0) |
| South Indian | 0% (0) | 6.3% (1) |
| RA characteristics |  |  |
| RA duration since diagnosis (median [IQR]) | 4.4 [0.3 – 11.1] years | N/A |
| RA duration since initial symptoms (median [IQR]) | 5.4 [1.9 – 14.5] years | N/A |
| Rheumatoid Factor positive (%, n) | 100% (16) | N/A |
| Anti-CCP antibody positive (%, n) | 93.8% (15) | N/A |
| Treatment naïve (%, n) | 18.8% (3) | N/A |
| TNF inhibitor naïve (%, n) | 62.5% (10) | N/A |

**Supplementary Table 1**. Characteristics of RA patients and controls. *Totals may not add up to 100% due to rounding*

| Antibody (clone) | Fluorochrome | Vendor | Isotype | Catalog no. |
| --- | --- | --- | --- | --- |
| HLA-DR (G46-6) | V500 | BD Biosciences | Mouse IgG2a, κ | 561224 |
| CD16 (3G8) | BUV-496 | BD Biosciences | Mouse IgG1, κ | 612944 |
| CD14 (63D3) | SB550 | Biolegend | Mouse IgG1, κ | 367148 |
| CD56 (NCAM16.2) | BUV737 | BD Biosciences | Mouse IgG1, κ | 748609 |
| CD141 (1A4) | BB515 | BD Biosciences | Mouse IgG1, κ | 565084 |
| CD1c (L161) | AF647 | Biolegend | Mouse IgG1, κ | 331510 |
| CD123 (6H6) | BV510 | BD Biosciences | Mouse IgG1, κ | 751831 |
| CD303 (201A) | APCFire750 | Biolegend | Mouse IgG2a, κ | 354236 |
| CD45RA (HI100) | BUV395 | BD Biosciences | Mouse IgG2b, κ | 740298 |
| CCR2 (K036C2) | PE-Cy7 | Biolegend | Mouse IgG2a, κ | 357212 |
| CD11c (3.9) | eFluor450 | Invitrogen | Mouse IgG1, κ | 48-0116-42 |
| CD45 (2D1) | PerCP | Biolegend | Mouse IgG1, κ | 368506 |
| CD83 (HB15e) | PE-Cy5 | Biolegend | Mouse IgG1, κ | 305310 |
| CD86 (IT2.2) | BV711 | Biolegend | Mouse IgG2b, κ | 305440 |
| CCR7 (G043H7) | BV421 | Biolegend | Mouse IgG2a, κ | 353208 |
| CD155 (PVR) (SKII.4) | PE-Dazzle594 | Biolegend | Mouse IgG1, κ | 337616 |
| CD275 (ICOS-L) (2D3) | PE | Biolegend | Mouse IgG2b, κ | 309404 |
| CD163 (GHI/61) | BV785 | Biolegend | Mouse IgG1, κ | 333632 |
| CD15 (W6D3) | BV605 | BD Biosciences | Mouse IgG1, κ | 663987 |
| CCR3 (5E8) | BUV805 | BD Biosciences | Mouse IgG2b, κ | 749025 |
| CD3 (SK7) | PerCP5.5 | Biolegend | Mouse IgG1, κ | 344808 |
| CD19 (HIB19) | Spark NIR 685 | Biolegend | Mouse IgG1, κ | 302270 |
| Viability | Propidium iodide | Sigma-Aldrich | N/A | 537059 |
|  | Live/Dead Blue | ThermoFisher | N/A | L23105 |

**Supplementary Table 2** Antibodies and Fluorochromes used for spectral flow cytometry.

| **DC2** (CD1c^+^) |  | CD56  BUV737 | CD163  BV785 | CCR2  PE-Cy7-A | CCR3  BUV805 | CCR7  BV421 | CD86  BV711 | CD14  SB550 | CD275  PE | HLA-DR  V500 | CD45RA  BUV395 |
| --- | --- | --- | --- | --- | --- | --- | --- | --- | --- | --- | --- |
| RA (n=16) |  | 9150  (8124) | 6668  (1533) | 220716  (39129) | 3815  (1794) | 3535  (8124) | 5579  (1707) | 10490  (3316) | 3701  (2016) | 229027  (30002) | 55233  (23308) |
| HC (n=16) |  | 3852  (1877) | 5520  (1517) | 217396  (42756) | 3379  (1346) | 1102  (1478) | 3930  (1335) | 8167  (2877) | 2197  (1297) | 206766  (40330) | 44034  (12426) |
| p-Value |  | 0.0164 | 0.0416 | 0.8203 | 0.5438 | 0.0002 | 0.0048 | 0.0427 | 0.0178 | 0.0866 | 0.1002 |

**Supplementary Table 3** DC2 (CD1c+) phenotypes. Mean Fluorescence Intensities (SD) of in rheumatoid arthritis (RA) patients and healthy donors measured by spectral cytometry. HD: Healthy Donor. N.S.: Not significant.

| **DC1** (CD141^+^) | CD56  BUV737 | **CCR2** PE-Cy7-A | CCR7  BV421 | **CD86** BV711 | **CD14** SB550 | CD275  PE | HLA-DR  V500 | CD45RA  BUV395 |
| --- | --- | --- | --- | --- | --- | --- | --- | --- |
| RA (n=16) | **13.8K** (14.8K) | **55.9K** (18.0K) | **3582** (2005) | **3308** (1301) | **3397** (16.1K) | 4660  (2087) | 168K  (47.9K) | 30.9K  (18.6K) |
| HC (n=16) | **5913** (5725) | **56.6K** (18.0K) | **1142** (1846) | 3232  (950) | **9028** (14.5K) | 3085  (2133) | 150K  (29.7K) | 30.4K  (20.8K) |
| p-Value | N.S. | N.S. | 0.0012 | N.S. | N.S. | 0.0433 | N.S. | N.S. |

**Supplementary Table 4** DC1 (CD141+). Mean Fluorescence Intensities (SD) of in rheumatoid arthritis (RA) patients and healthy controls was measured by spectral cytometry. Not differing significantly: CCR3, CD11c eFluor450, CD163 BV785, CD141 BB515, CD14 SB550, CD45 PerCP, CD15

| **pDC** (CD303^+^CD123^+^) | CD56  BUV737 | **CCR2** PE-Cy7-A | CCR7  BV421 | **CD83** PE-Cy5 | **CD86** BV711 | **CD14** SB550 | CD275  PE | HLA-DR  V500 | **CD11c** eFluor 450 | CD45RA  BUV395 | CD123  BV510 | CD303  APC-Fire750 |
| --- | --- | --- | --- | --- | --- | --- | --- | --- | --- | --- | --- | --- |
| RA (n=16) | **6721** (3501) | 181K  (50.6K) | 4344  (1071) | 1011  (2376) | 3379  (1216) | **5057** (3645) | **2813** (1353) | **125K** (26.3K) | -1572  (2794) | **181K** (23.4K) | 65.9K  (19.3K) | 50.5K  (11.1K) |
| HC (n=16) | **4329** (981) | 210K  (46.8K) | 3669  (1338) | -613  (1982) | 2862  (972) | 4982  (3451) | **1974** (1202) | **118K** (25.0K) | **-1645** (981) | **187K** (20.8K) | 55.8K  (22.2K) | 49.3K  (11.2K) |
| p-Value | 0.0133 | N.S. | N.S. | 0.0443 | N.S. | N.S. | N.S. | N.S. | N.S. | N.S. | N.S. | N.S. |

**Supplementary Table 5** Phenotype of Plasmacytoid Dendritic Cells (pDC) in RA and healthy controls.

Mean Fluorescence Intensities (SD) of in rheumatoid arthritis (RA) patients and healthy donors measured by spectral cytometry. Not differing significantly: CD11c eFluor 450, CD163 BV785, CD155 PE-Dazzle594. HC: Healthy control. N.S.: Not significant

|  | RA Patient 1 | RA Patient 2 | RA Patient 3 |
| --- | --- | --- | --- |
| Patient ID | 005 | 009 | 013 |
| Age | 37 | 59 | 78 |
| Gender | Female | Male | Female |
| Ethnicity | White | White | White |
| Rheumatoid Factor | 74 IU/ml | 197 IU/ml | negative |
| CCP | > 250 U/ml | > 250 U/ml | 125 U/ml |
| Smoking status | never | smoker | never |
| Periodontal disease | No | No | No |
| Symptomatic Joints | Bilateral ankles and knees, bilateral wrists, MCP and PIP joints | Bilateral feet, bilateral ankles, bilateral knees, bilateral wrists, all MCP and PIP joints | Bilateral wrists, all MCP and PIP joints |
| RA treatment | Oral methotrexate, prednisone taper | Oral methotrexate, prednisone taper | Oral methotrexate, prednisone taper |
| Initial CRP | 28.5 mg/L | 77.2 mg/L | 52.9 mg/L |
| Initial ESR | 50 mm/hr | > 120 mm/hr | 40 mm/hr |
| RA symptom Duration | 2 months | 3 years | 3 months |
| Time since RA diagnosis | 1 week | 1 week | 1 week |

**Supplementary Table 6** Clinical characteristics of Rheumatoid Arthritis Index Patients

|  | RA  (n=16) | RA ‘index’ patients  (n=3) | Non-index RA patients  (n=13) |
| --- | --- | --- | --- |
| Age (mean, SD) | 54.6 ± 16.2 years | 59.0 ± 20.5 years | 53.8 ± 15.9 years |
| Gender (%, n) |  |  |  |
| Female | 75% (12) | 67% (2) | 77% (10) |
| Male | 25% (4) | 33% (1) | 23% (3) |
| Other | 0% (0) | 0% (0) | 0% (0) |
| Ethnicity (%, n) |  |  |  |
| White | 75% (12) | 100% (3) | 69% (9) |
| Black | 12.5% (2) | 0% (0) | 15.5% (2) |
| Native American | 12.5% (2) | 0% (0) | 15.5% (2) |
| South Indian | 0% (0) | 0% (0) | 0% (0) |
| RA characteristics |  |  |  |
| RA duration since diagnosis (median [IQR]) | 4.4 [0.3 – 11.1] years | 0.1 [0.04 – 0.4] years | 6.1 [3.49 – 13.3] years |
| RA duration since initial symptoms (median [IQR]) | 5.4 [1.9 – 14.5] years | 0.9 [0.24 – 3.1] years | 6.8 [2.45 – 16.1] years |
| Rheumatoid Factor positive (%, n) | 100% (16) | 100% (3) | 100% (13) |
| Anti-CCP antibody positive (%, n) | 93.8% (15) | 100% (3) | 92.3% (12) |
| Treatment naïve (%, n) | 18.8% (3) | 100% (3) | 0% (0) |
| TNF inhibitor naïve (%, n) | 62.5% (10) | 100% (3) | 53.8% (7) |

**Supplementary Table 7** Summary of index and non-index RA patients.

|  | **HC** (n=16) | **RA** (n=16) | p-value |
| --- | --- | --- | --- |
| Classical  (CD14^hi^CD16^-^) | 79.3%  IQR [74.67-85.65] | 75.5%  IQR [64.14-81.81] | N.S. |
| Nonclassical  (CD14^lo^CD16^+^) | 3.38%  IQR [1.37-5.21] | 4.87%  IQR [2.45-9.82] | N.S. |
| Intermediate  (CD14^hi^CD16^+^) | 4.74%  IQR [1.21-6.64] | 14.25%  IQR [7.70-17.98] | 0.0037 |

**Supplementary Table 8**. Median of monocyte subsets in rheumatoid arthritis and healthy donors (in % of non-lymphoid DR^hi^APC).
HD: Healthy Donor RA: Rheumatoid Arthritis. N.S.: Not significant. IQR: Interquartile

**Supplemental Data of APC (HLA-DR+) populations and relevant Jupyter/Python code:**

<https://github.com/christian-geier/apc-data>

**Supplemental method information**

As suggested by Lee JA, Spidlen J, Boyce K, Cai J, Crosbie N, Dalphin M, et al. MIFlowCyt: The Minimum Information about a Flow Cytometry Experiment. Cytometry A. 2008 Oct;73(10):926–30.

<https://www.ncbi.nlm.nih.gov/pmc/articles/PMC2773297/>

**Experiment overview**

1.1. **Purpose**

The purpose of the experiments was to determine the composition of blood antigen-presenting cells (HLA-DR+) from rheumatoid arthritis patients (RA) compared with healthy control donors (HC), by spectral cytometric analysis with an appropriate combination of lineage specific antibodies.

1.2. **Keywords**

Blood, rheumatoid arthritis, autoimmunity, antigen-presenting cells, dendritic cells, leukocytes, monocytes, granulocytes

1.3. **Experiment Variables**

Peripheral blood mononuclear cells (PBMC) from seropositive RA patients and HC donors (n=16 each group) were collected through standard venipuncture for cytometry. No interventions were performed.

1.4. **Organization**
     1.4.1. Name: SUNY Upstate Medical University
     1.4.2. Address: 766 Irving Avenue, Syracuse, NY, 13210, United States
1.5. **Primary Contact**
     1.5.1. Name: Christian Geier
     1.5.2.
1.6. **Dates**

Sample collection dates: November 17, 2020 – May 18, 2023

Sample processing dates: April 16, 2021 – October 17, 2023

1.7. **Conclusions**
- CD1c+ dendritic cells (DC2) are decreased in RA
- CD15+CD16+ cells, a subset staining positive for HLA-DR, appear in RA
  1.8. **Quality Control Measures**

PBMC collected from patients and appropriate healthy (no history of RA or other autoimmune conditions) controls, matched, by gender and age (within 10 years), with healthy control donors. Use of technical reference controls.

1.9. **Other Relevant Experiment Information**

N/A

2. **Flow Sample/Specimen Material Description**
   2.1. **Sample/Specimen material description**
        2.1.1. Biological Samples
               2.1.1.1. *Biological Sample Description*: 10-15 ml patient or healthy control donor blood, collected in heparinized tubes
               2.1.1.2. *Biological Sample Source Description*: Human (Homo sapiens) with rheumatoid arthritis, or healthy control
               2.1.1.3. *Biological Sample Source Organism Description:*
               2.1.1.4. Age.

Supplementary Table 1 contains demographic information for RA and HC

2.1.1.5. Gender

Supplementary Table 1 contains demographic information for RA and HC

2.1.2. Environmental Samples N/A
2.1.3. Other Samples N/A
2.2. **Sample Characteristics**

Expected/analyzed types of cells: lymphoid and myeloid populations, other low-density cells contained in the PBMC layer after Ficoll separation

2.3. **Sample Treatment Description**

See main method section for PBMC isolation and processing.

2.4. **Fluorescence Reagent Description**

Each sample has been stained according to Supplementary Table 2 and acquired as an unstained control for autofluorescence subtraction. Single stained controls with 1 or 2 uL of antibody/fluorochrome conjugates were used as described in the main method section.

3. **Instrument Details**
   3.1. **Instrument Manufacturer**

Cytek Biosciences, <www.cytekbio.com>

3.2. **Instrument Model**

Cytek Aurora™, 5 Laser Spectral Cytometer

Technical Specification at: <https://cytek-web.s3.amazonaws.com/cytekbio.com/documentation-center/Brochures/N9-20001_Cytek_Aurora_Brochure.pdf>

3.3. **Instrument Configuration and Settings**
     3.3.1. **Flow Cell and Fluidics**

The instrument has not been altered; medium flow rate 30 µL/min

3.3.2. **Light Sources**

The instrument has not been altered; five laser configuration

355 nm: 20 mW

405 nm: 100 mW

488 nm: 50 mW

561 nm: 50 mW

640 nm: 80 mW

3.3.3. **Excitation Optics Configuration**

The instrument has not been altered; Flat-Top laser beam profile with narrow vertical beam height.

3.3.4. **Optical Filters**

The instrument has not been altered

3.3.5. **Optical Detectors**

The instrument has not been altered; five avalanche-photodiodes (APD) detector arrays. Detector voltages were set according to manufacturer recommendations.

Violet detector module: 16 channels unevenly spaced (420-829 nm)

Blue detector module: 14 channels unevenly spaced (498-829 nm)

Red detector module: 8 channels unevenly spaced (652-829 nm)

Yellow-Green detector module: 10 channels unevenly spaced (567-829 nm)

Ultraviolet detector module: 16 channels unevenly spaced (365-829 nm)

3.3.6. **Optical Paths**

The instrument has not been altered; Laser delays are automatically adjusted during instrument QC.

3.4. **Other Relevant Instrument Details**

[Aurora User Guide](https://welcome.cytekbio.com/hubfs/N9-20006%20Rev.%20E_Cytek%20Aurora%20User%20Guide.pdf)

4. **Data Analysis Details**
   4.1. **List-mode Data Files**

FCS Files are available from the primary contact. Python/Jupyter scripts used have been deposited and can be accessed in the following repository: <https://github.com/christian-geier/apc-data>

4.2. **Similarity matrix of fluorochromes used**

**Complexity index: 19.7**

4.3. **Data Transformation Details**
     4.3.1. Purpose of Data Transformation

Visualization and gating

4.3.2. Data Transformation Description

SpectroFlo default visualization settings have been used for gating:

Scaling: Biexponential scaling with auto, except linear scaling for FSC and SSC and time

4.3.3. Other Relevant Data Transformation Details

We used t-distributed stochastic neighbor embedding (t-SNE) with parameters listed in the main method section.

4.4. **Gating (Data Filtering) Details**

The same gating strategy has been used for all data files (Supplementary Figures 1 and 2)

4.4.1. Gate Descriptions
- For reference APC populations (Supplementary Figure 1)
  We gated large live singlet CD45+ leukocytes (A-D). Lymphoid cells (CD3+, CD19+) and HLA-DRneg/lo cells were gated out (E, F). Classical, intermediate, and non-classical monocytes were gated by CD14 and CD16 expression (G). We then gated pDC as CD303+CD123+ population (H). After applying a NOT(pDC) gate (that removes pDC gated in H), we gated CD141+ DC1 (blue events) and CD1c+ DC2 (orange events)
- For CD15^+^CD16^+^ and CD15^+^CD16^+^DR^hi^ (Supplementary Figure 2)
  We gated live singlet CD45+ leukocytes (A-C). The “large leukocytes” gate was widened to ensure full capture of granulocytic populations (D, red gate and dashed ellipse). Lymphoid cells (CD3+, CD19+) were gated out (E). CD15+CD16+ cells were gated (F). Separately, large non-T&B cells shown in E were gated for HLA-DRhi, HLA-DRneg/lo cells were gated out (G). The CD15+CD16+ DRhi population was gated (H). (E, F).
  4.4.2. Gate Statistics
         Displayed in Figures 2 and 5 and Supplementary Figures and Tables. Gate statistics have been deposited and can be accessed in the following repository: <https://github.com/christian-geier/apc-data>
  4.4.3. Gate Boundaries

Displayed in Supplementary Figures 1 and 2. DR^hi^ was set as greater than the mean value of the DR+ population.

4.4.4. Other Relevant Gate Information

For CD15^+^CD16^+^ and CD15^+^CD16^+^DR^hi^, the leukocyte gate was extended (Supplementary Figure 2) to allow for the inclusion of granular populations.
